# Similar Wires, Different Fires: Reconsidering the Link Between Resting-state Functional Connectivity and Psychopathic Traits

**DOI:** 10.64898/2026.01.12.26343689

**Authors:** Sofiia Tatosh, Stephane A. De Brito, Sally Chester, Tatsuyoshi Ogawa, Maki Terao, Ryusuke Nakai, Nobuhito Abe, Jules R. Dugré

## Abstract

Psychopathy is a multifaceted construct encompassing affective, interpersonal, behavioural, and antisocial traits. Intrinsic brain organization, captured via functional connectivity, is widely used to map brain-behaviour relationships and to characterize the neural underpinnings of psychopathology. Prior work reports associations between psychopathic traits and altered connectivity in the frontoparietal control, default mode, and salience networks. However, current evidence is constrained by small sample sizes and the reliance on Western, Educated, Industrialized, Rich, and Democratic populations. Therefore, cross-cultural predictive modeling is crucial for identifying robust and generalizable biomarkers linked to psychopathic traits.

To address these gaps, we tested whether resting-state functional connectivity (rsFC) predicts psychopathic traits in a non-Western community cohort. Participants (n=97, 52 females, community sample, Japan) completed the Self-Reported Psychopathy Scale-Short Form (SRP-SF) and underwent structural and resting-state functional magnetic resonance imaging. Connectome-based Predictive Modeling (CPM) with cross-validation was applied, and generalizability was evaluated in an independent dataset (n=107, 68 females, community sample, Germany).

Across both cohorts, CPM failed to predict psychopathy scores out-of-sample, regardless of model parameters, psychopathic subdimensions, or potential sex differences. Post-hoc analyses indicated, however, that individuals with higher psychopathy scores exhibited more homogeneous rsFC patterns, whereas those with lower scores showed greater variability, suggesting score-dependent heterogeneity in brain-behaviour mapping. These findings highlight key challenges for predictive modeling and point to several future directions, including larger samples, the use of non-linear models, and potentially a shift toward individual-level connectome characterization to better capture heterogeneous neural substrates of similar behavioural phenotypes.

## 2 Introduction

Psychopathy is a personality disorder characterized by a set of affective, interpersonal, behavioral, and antisocial traits such as shallow affect, lack of empathy, grandiosity, impulsivity, manipulativeness, poor decision-making, parasitic lifestyle, and disregard for social norms, including criminal versatility[1]. Factor analyses of the Hare Psychopathy Checklist measures (PCL/PCL-R)[1, 2], widely used to operationalize psychopathy in both clinical and offender populations, have identified two broad trait dimensions[3–6]: Factor 1, indicating profound emotional blunting and tactical exploitation of others; and Factor 2, reflecting a predisposition toward impulsive behavior and chronic rule-breaking. This two-factor model is also frequently described at the level of four facets: Interpersonal and Affective facets under Factor 1, and Lifestyle and Antisocial facets under Factor 2[1, 5, 6].

While often associated with criminal and violent behaviour (10-25% of individuals in forensic samples exhibit elevated psychopathy levels[7], [8], [9]), individuals with psychopathy can be found within the community (e.g., corporate, military, political, and academic sectors) with an estimated prevalence around 2% [10]. Psychopathy is thought to result from a complex interplay between genetic and environmental factors[11], with growing evidence that psychopathy is associated with structural and functional brain abnormalities[12], [13]. Indeed, converging evidence from task-based functional magnetic resonance imaging (MRI), lesion, and structural MRI studies has consistently shown involvement of a network of cortical (e.g., orbitofrontal, anterior cingulate, insular, and temporal cortices) and subcortical (e.g., amygdala, striatum) regions in the pathophysiology of psychopathy[11], [14], [15]. In particular, studies that have employed empathy-related fMRI tasks have revealed hypoactivation of key affective regions - amygdala, ventromedial prefrontal cortex (vmPFC), and anterior insula - in individuals scoring high on psychopathic traits, whereas decision-making tasks show attenuated striatal responses during reward anticipation[16].

Beyond task-evoked activation, resting-state functional MRI (rs-fMRI) has been widely used as an alternative method to study psychiatric disorders[17]. Rs-fMRI measures spontaneous low-frequency fluctuations in the blood oxygen-level dependent (BOLD) signals at rest, enabling the assessment of functional connectivity (rsFC) as the temporal correlation among these BOLD time series across anatomically distinct regions[17]. The absence of explicit task requirements yields high test-retest reliability. Hence, canonical resting-state networks (RSNs) emerge consistently across individuals, underpinning core cognitive and affective processes: the default mode network, involved in self-referential processing and social cognition; the salience network, implicated in responding to salient stimuli; the frontoparietal network, linked to goal-directed control processes; and primary sensorimotor network, engaged in maintaining basic perceptual and motor responses[18]. Current views in psychiatry hold that psychiatric conditions - including psychopathy[19], [20] - stem from dysfunctions in large-scale brain network communication rather than from lesions in a single localized region[21]. Despite some inconsistencies across studies, these connectivity disruptions converge on three large-scale networks - the default mode network (DMN), frontoparietal control network (FPN), and salience network (SN) - as fundamental to psychopathy[22], [23], [24]. Of note, limited evidence suggests similar alterations across age and sex[24], although no formal statistical comparisons of sex differences have been conducted to date.

Given the promise of connectivity-based biomarkers for personalized diagnosis and intervention, there has been a growing emphasis on developing tools to predict psychopathy severity at the individual level[25]. Among these methods, connectome-based predictive modelling (CPM) has gained particular prominence for its ability to predict behavioral phenotypes from whole-brain connectivity[26]. In contrast to simple mass-univariate analyses, which test connection-behavior association independently, CPM jointly considers the predictive contribution of a set of connections, yielding greater sensitivity to subtle, multivariate brain-behavior relationships and enhanced out-of-sample generalizability[26]. Indeed, CPM ipsychotic treatment response in first-episode psychosis[27], suicide risk in late-life depression[28], and compulsion severity in obsessive-compulsive disorder[29]. In a recent study using a sample of 84 college students, CPM was applied to whole-brain rsFC and showed that the set of connections negatively correlated with Levenson Self-Report Psychopathy (LSRP) scores significantly predicted both total LSRP and secondary psychopathy scores (but not primary psychopathy); this predictive network was anchored by high-degree nodes in the anterior prefrontal cortex, orbitofrontal cortex, anterior cingulate cortex, and insula, and notably, connectivity between the occipital network and the cingulo-opercular network emerged as the strongest predictor of individual differences in psychopathy severity[30].

Despite a growing body of group-level functional findings in psychopathy, clinical translation remains limited - most studies do not evaluate individual-level prediction, rely on small, unrepresentative cohorts, and often omit key validation steps[31]. Additionally, the overwhelming majority of human neuroimaging research relies on participants from Western, Educated, Industrialized, Rich, and Democratic (WEIRD) societies. Indeed, 90% of published psychology studies sample exclusively from WEIRD populations[32], [33], [34]. In sum, the field suffers from a“replication crisis” that is exacerbated by an increasing reliance on computational modelling and predictive analytics applied to large-scale, WEIRD-dominated datasets[35].

To address these gaps, the present study applied a connectome-based predictive modelling (CPM) framework to whole-brain resting-state functional connectivity (rsFC) data in a community sample of Japanese adults. Our primary goal is to predict individual scores on the Self-Report Psychopathy Scale-Short Form (SRP-SF) total score and factors 1 (interpersonal/affective) and 2 (lifestyle/antisocial) from intrinsic connectivity patterns. We hypothesized that functional connectivity edges within the salience network, default mode network, frontoparietal network, and subcortical regions would predict variance in psychopathic traits severity in a Japanese community sample. To test the replicability of our findings, we then applied our model directly to another sample, including Western participants, the MPI-Leipzig Mind-Brain-Body dataset[36], to assess whether the same network edges would predict psychopathy scores with comparable accuracy. Finally, we enhanced the standard CPM pipeline by (1) weighting each predictive edge to amplify the influence of the most relevant connections in the network summary, and (2) using the Network-Based Statistic[37] for feature selection, which identifies suprathreshold connected subnetworks and has been shown to outperform univariate thresholding and other pre-existing feature selection algorithms[38]. By integrating these methodological refinements with cross-cultural replication in a German validation sample, this study is the first to test the generalizability of resting-state connectivity patterns associated with psychopathy scores across populations from different cultures.

### Aims and Hypotheses

Based on established literature, we formulated the following research questions and predictions:

**Research question 1.** Can resting-state functional connectivity serve as a predictive marker of individual differences in psychopathy (Total score, Factor 1, Factor 2) within a Japanese community sample?

- **Hypothesis 1.1.** Whole-brain resting-state functional connectivity patterns will predict Total, Factor 1, and Factor 2 psychopathy scores in the Japanese community sample.
**Research question 2.** Which resting-state networks contribute most significantly to the prediction of psychopathy dimensions?

- **Hypothesis 2.1.** Higher psychopathy scores will be associated with stronger disconnection within the default mode network (i.e., medial prefrontal cortex, posterior cingulate), salience network (i.e., insula, anterior cingulate cortex), frontoparietal network (i.e., dorsolateral prefrontal cortex), and subcortical regions (i.e., amygdala, striatum).
**Research question 3.** Does a connectivity-based predictive model derived from the Japanese community sample generalize, without retraining, to predict the severity of psychopathic traits in an independent cohort from a different culture?

- **Hypothesis 3.1.** The model will generalize to the independent cohort, yielding significant, but likely attenuated, prediction of psychopathy scores relative to the original sample.

#### Exploratory analyses

- Does weighted-sum CPM outperform unweighted CPM in cross-validated prediction of Total, Factor 1, and Factor 2?
- Does an NBS-derived subnetwork for feature selection yield more accurate cross-validated predictions of Total, Factor 1, and Factor 2 scores than conventional CPM-selected edges?
- Are there any sex differences in (a) the accuracy of CPM-based psychopathy predictions and (b) the specific connectivity patterns driving those predictions?

*Due to the lack of prior research, these analyses are exploratory, and no a priori hypotheses were formulated*.

## 3 Methods

### 3.1 Data Preparation

To investigate the relationship between brain function and psychopathic traits, and to test whether any brain-behavior associations would generalize across samples, we utilized two independently collected resting-state fMRI datasets. These comprised a cohort from Japan and a cohort from Germany. This section describes the demographic and acquisition details for each dataset, along with the standardized preprocessing pipeline applied to both cohorts to ensure data comparability.

#### 3.1.1 Japanese dataset

The primary dataset is a cohort of structural and resting-state functional MRI scans collected from a Japanese community sample. This cohort comprises 97 healthy adult participants (right-handed, mean age = 26.9 ± 5.33, 52 females). One participant was excluded due to a missing fMRI scan, resulting in a total of 96 subjects analysed. Psychometric assessments and neuroimaging were conducted at Kyoto University, Japan. Psychopathic traits were measured via the Self-Reported Psychopathy Scale Fourth Edition (SRP-4) Short Form[39]. Participant’s responses on the SRP-SF were aggregated to derive a total psychopathy score and four facet subscale scores: antisocial behaviour (ASB) (8 items, *α* = 0.63), e.g., “I have assaulted a law enforcer or social worker”; callous affect (CA) (7 items, *α* = 0.67), e.g., “I never feel guilty when I hurt people”; interpersonal manipulation (IM)(7 items, *α* = 0.68), e.g., “I like pushing people to their breaking point”; and erratic lifestyle (EL)(7 items, *α* = 0.82), e.g., “I like having sex with people I barely know”. For the present analyses, in line with established guidelines[40], we also computed a Factor 1 score by summing the CA and IM subscale responses (14 items, *α* = 0.85), and a Factor 2 score by summing the ASB and EL subscale responses (15 items, *α* = 0.76). All participants provided informed consent in accordance with the guidelines of the Ethics Committee at Kyoto University.

All magnetic resonance imaging was performed at Kyoto University on a Siemens 3T Verio scanner equipped with a 32-channel head coil. A high-resolution T1-weighted structural image was acquired using a Magnetization Prepared Rapid Acquisition Gradient Echo (MPRAGE) sequence. Acquisition parameters were: repetition time (TR) = 2500 ms, echo time (TE) = 2.18 ms, flip angle = 8^◦^, field of view (FOV) = 256 mm, and voxel size = 0.8*x*0.8*x*0.8 mm, yielding 224 sagittal slices. The total scan duration was 5 minutes and 22 seconds. fMRI images were acquired using an echo-planar imaging (EPI) sequence covering the entire brain. Acquisition parameters were: TR = 1300 ms, TE = 40 ms, flip angle = 66^◦^, FOV = 24 cm, and voxel size = 2.5*x*2.5*x*2.5 mm. A total of 60 axial slices were acquired. The functional scan lasted 10 minutes.

#### 3.1.2 German dataset

The second dataset was sourced from MPI-Leipzig Mind-Brain-Body open-access database[36]. The initial available cohort included 107 healthy participants (age range = 20–80; 68 females). However, to ensure demographic comparability with the Japanese dataset, this sample was later restricted to match the age range of the primary cohort, resulting in a total of 87 participants (age range = 20–35; 48 females). Psychometric assessments and neuroimaging were conducted at the Max Planck Institute for Human Cognitive Neurology at the University of Leipzig, Germany, as part of the MPI-Leipzig Mind-Brain-Body functional connectome phenotyping project[41]. Participants completed the Short Dark Triad (SD3) in addition to other personality measures. In the present analyses, the psychopathy SD3 subscale (e.g., “It’s true that I can be mean to others”) was used as the target variable for testing hypotheses. All participants provided written informed consent, and the study was approved by the ethics committee at the medical faculty of the University of Leipzig.

High-resolution T1-weighted structural images were acquired on a 3T Siemens Magnetom Verio scanner using a 3D Magnetization-Prepared 2 Rapid Acquisition Gradient Echos (MP2RAGE) sequence. Acquisition parameters were: TR = 5000 ms, TE = 2.92 ms, flip angles = 4^◦^ and 5^◦^, FOV = 256*x*240*x*176 mm, and voxel size = 1.0*x*1.0*x*1.0 mm, resulting in a single 3D volume with 176 sagittal slices. Resting-state fMRI images were acquired using a T2*-weighted gradient-echo EPI sequence. Acquisition parameters: TR = 1400 ms, TE = 39.4 ms, flip angle = 69, FOV = 202*x*202 mm^2^, voxel size = 2.3*x*2.3*x*2.3 mm, and 64 axial slices. Each run consisted of 657 volumes and lasted 15 minutes and 30 seconds.

#### 3.1.3 Data preprocessing (CONN toolbox)

All rs-fMRI data from both cohorts were preprocessed using the CONN (22.v2407) functional connectivity toolbox[42]. The default preprocessing pipeline was employed. Specifically, functional and anatomical data were preprocessed using a modular preprocessing pipeline[43] including realignment with correction of susceptibility distortion interactions, slice timing correction, outlier detection, direct segmentation and MNI-space normalization, and smoothing. The functional data were realigned using SPM realign and unwarp procedure[44], where all scans were coregistered to a reference image (first scan) using a least squares approach and a 6 parameter (rigid body) transformation[45], and resampled using b-spline interpolation to correct for motion and magnetic susceptibility interactions. Temporal misalignment between different slices of the functional data (acquired in interleaved Siemens order) was corrected following SPM slice-timing correction (STC) procedure[46], [47], using sinc temporal interpolation to resample each slice BOLD timeseries to a common mid-acquisition time. Potential outlier scans were identified using ART[48] as acquisitions with framewise displacement above 0.9 mm or global BOLD signal changes above 5 standard deviations[49], [50], and a reference BOLD image was computed for each subject by averaging all scans excluding outliers. Functional and anatomical data were normalized into standard MNI space, segmented into grey matter, white matter, and CSF tissue classes, and resampled to 2 mm isotropic voxels following a direct normalization procedure[50], [51] using SPM unified segmentation and normalization algorithm[52], [53] with the default IXI-549 tissue probability map template. Last, functional data were smoothed using spatial convolution with a Gaussian kernel of 6 mm full width half maximum (FWHM)[54].

#### 3.1.4 Connectome Construction

Following data preprocessing, brain activity was first summarized into a set of functionally homogeneous regions (parcels) that served as nodes in the functional connectome. Cortical parcels were defined using the 400-region Schaefer atlas[55], supplemented by a standard set of subcortical[56] and cerebellar[57] ROIs, resulting in a total of *P* = 421 distinct nodes.

To map each individual’s brain organization at rest, functional connectivity was measured to reflect the co-activation between pairs of regions over time. First, the mean pre-processed BOLD time series was extracted from each ROI for every subject. These time courses were band-pass filtered between 0.008 and 0.09 Hz. Functional connectivity was defined as a measure of strength and direction of a linear relationship between two time series. This was achieved by computing the Pearson correlation coefficient (*r*) between every pair of ROI signals. Mathematically, Pearson correlation between two time series *x*(*t*) and *y*(*t*) is:

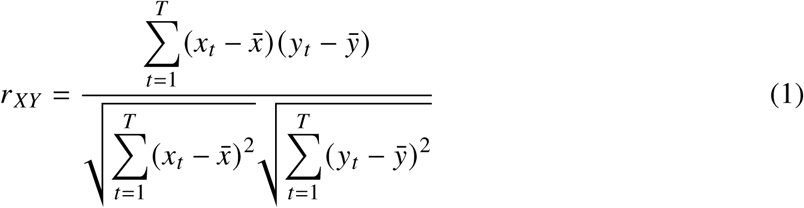

where *x*_*t*_ and *y*_*t*_ denote the BOLD signal at time point *t* in each region, *x̄* and *ȳ* are the mean signals across time points, and *T* is the number of time points in the time series. Applying pairwise Pearson correlation produced a *P* × *P* correlation matrix for each participant, where each entry represents the strength of a functional connection between two nodes. Finally, all correlation values were transformed using Fisher’s *z*-transformation to stabilize variance for subsequent analyses:

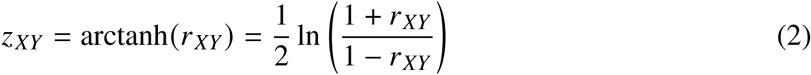

For sufficiently large *T*, the transformed values *z*_*XY*_ are approximately normally distributed with variance ≈ 1/(*T* − 3), which makes them more suitable for use in linear models. The resulting connectome provided a unique representation of each individual’s whole-brain functional organization, serving as the primary data for the predictive modelling described in the next section.

### 3.2 Connectome-Based Predictive Modelling

Connectome-based predictive modelling is a data-driven framework that uses whole-brain functional connectivity patterns to predict individual differences in behavior or clinical traits. This approach typically involves several key stages: (1) feature selection to identify trait-relevant connections, (2) feature summarization to create a single ‘network strength’ score per participant, (3) model building to relate network strength to the behavioral trait, and (4) a rigorous assessment of the model’s predictive significance, often using permutation testing[26].

After data preprocessing and connectome construction, the CPM was applied to achieve three primary objectives. First, to test whether individual differences in whole-brain functional connectivity could successfully predict psychopathic traits (Total, Factor 1, and Factor 2 scores). Second, to identify the specific neural circuits driving this prediction. And third, to assess whether the predictive model generalizes to a dataset with a different cultural background (see Hypotheses).

**Figure 1:**
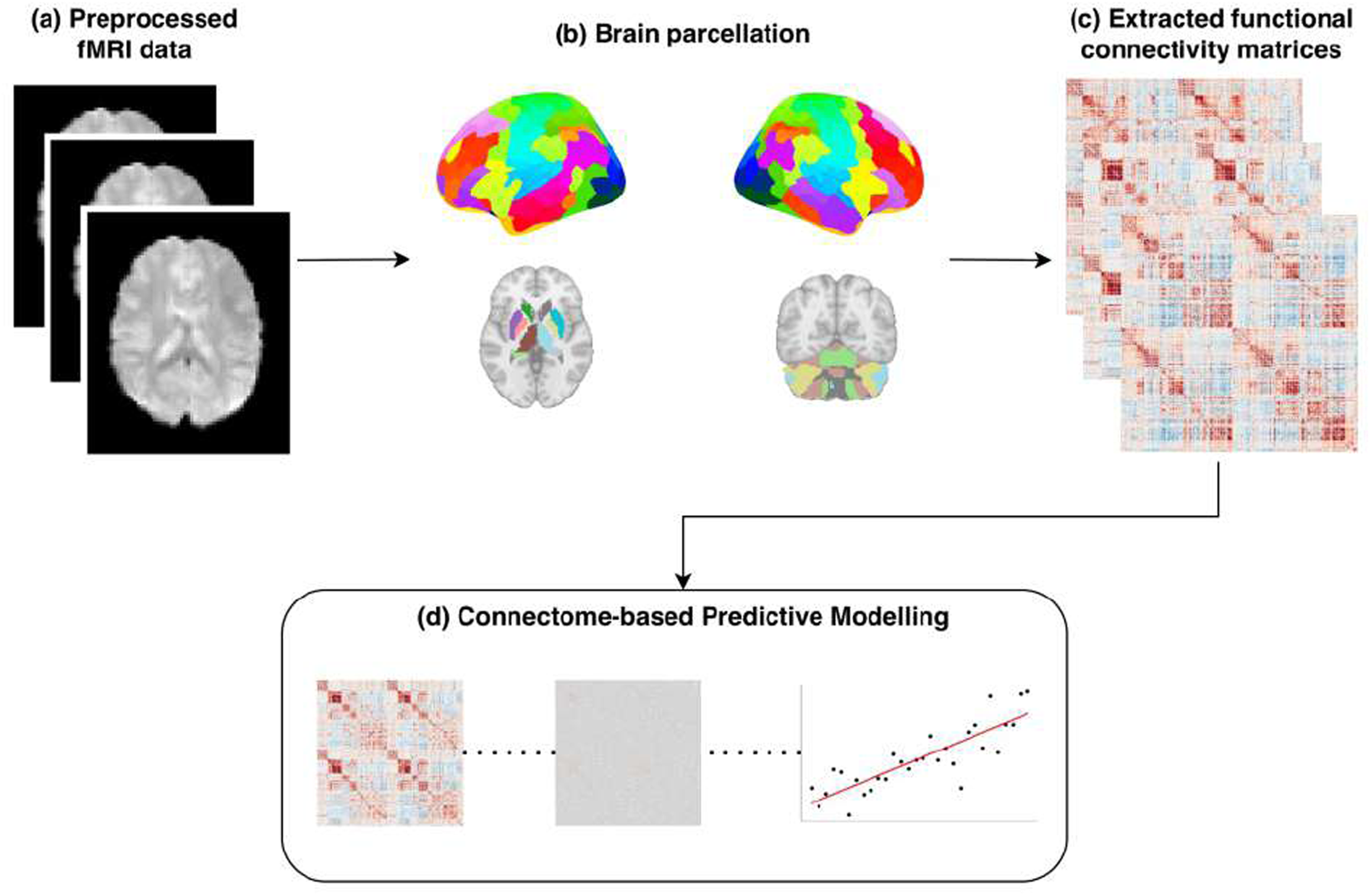
Overview of the analytical pipeline. (a) Preprocessed resting-state fMRI data were obtained for each participant; (b) data were parcellated using a standard cortical and subcortical atlas; (c) subject-specific functional connectivity matrices were extracted; (d) the matrices were used as an input to connectome-based predictive modeling to test whether whole-brain connectivity predicts individual differences in psychopathic traits.

**Figure 2:**
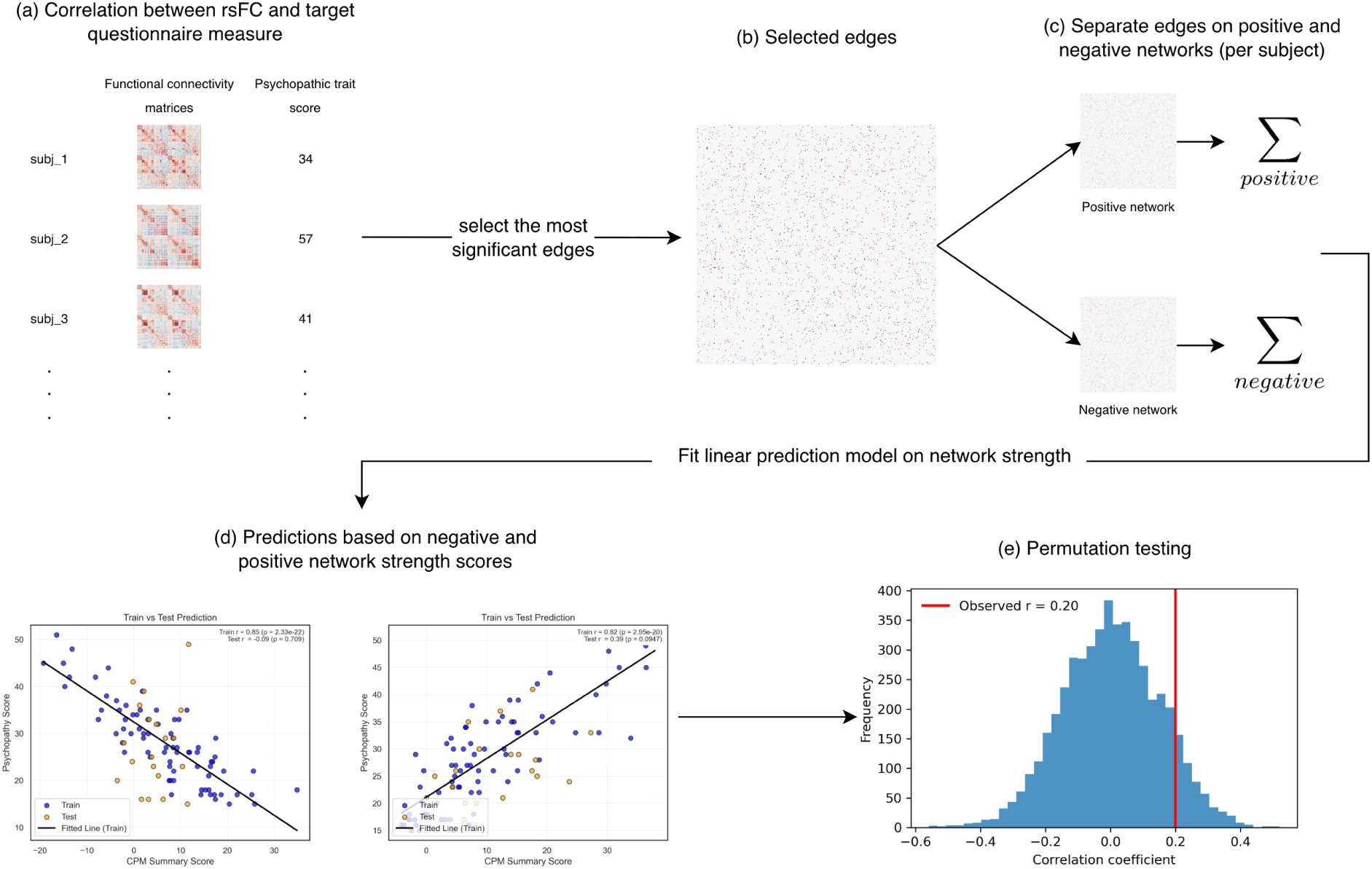
Overview of the connectome-based predictive modeling (CPM) pipeline. Schematic overview of the connectome-based predictive modeling (CPM) pipeline used in this study. (a) For each participant, whole-brain resting-state functional connectivity (rsFC) matrices are correlated with individual psychopathic trait scores at the edge level. (b) Edges that exceed a predefined significance threshold are selected. (c) Significant edges are divided into positive and negative networks, and for each subject network strength scores are computed by summing edge weights within each network. (d) Positive and negative network strength scores are entered into a linear model to predict psychopathy scores in the training set and generate out-of-sample predictions in the test set. (e) Model performance is assessed with permutation testing by shuffling psychopathy scores to build a null distribution of prediction coefficients and comparing it with the observed correlation.

Given the established success to predict a range of individual differences from whole-brain FC[58], [59], [60], [61], [62], CPM was selected as the primary tool to address the previously outlined hypotheses: (H1.1) whole-brain rsFC patterns will predict total, Factor 1, and Factor 2 psychopathy scores in the Japanese community sample, and (H3.1) a model derived in this sample will generalize, without retraining, to predict psychopathic traits in an independent German cohort.

Importantly, CPM yields not only prediction scores but also produces a set of predictive edges (functional connections whose strength reliably relates to the target trait). These edge sets can be projected onto canonical resting-state networks, allowing us to characterize which large-scale networks contribute most strongly to the predictions. Hence, CPM-derived edges allow us to address H2.1, which is whether higher psychopathy scores will be associated with stronger disconnection within the default mode, salience, frontoparietal networks, and subcortical regions.

#### 3.2.1 Baseline CPM implementation

To test Hypotheses 1.1 and 3.1, subject-wise resting-state functional connectivity (rsFC) matrices as predictors and psychopathy scores as target variables were used. For each participant *i* = 1,…, *N*, rsFC was summarized as a symmetric matrix *C*_*i*_ ∈ ℝ^*P*×*P*^, where *P* = 421 is the number of cortical, subcortical, and cerebellum parcels. Each entry *c*_*i*_ (*p*, *q*) is the Fisher-*z* transformed Pearson correlation between the preprocessed BOLD time series of parcels *p* and *q*. Since connectivity matrices are symmetrical, the duplicate values are removed by vectorizing the upper triangle of each matrix *C*_*i*_ (excluding the diagonal) into a connectivity vector **x**_*i*_ ∈ ℝ^*E*^, where *E* = *P*(*P* − 1)/2 is the number of resulting edges. Stacking these vectors yielded an *N* × *E* feature matrix

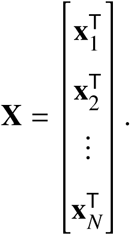

**x**_·*e*_ denotes the N-dimensional column vector containing the connectivity of edge *e* across all subjects:

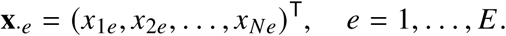

Psychopathic traits were represented by a behavioral vector **y** = (*y*_1_,…, *y*_*N*_)^T^ where *y*_*i*_ denotes the score of subject *i* on the relevant dimension (Total, Factor 1, or Factor 2; additional facet scores were considered in exploratory analyses).

To identify edges that are informative for predicting psychopathic traits (H1.1, H3.1), CPM performs feature selection separately within each training fold. For each edge *e* = 1,…, *E*, the Pearson correlation is computed between its connectivity strength across subjects **x**_·*e*_, and the corresponding psychopathy scores *y*: *r*_*e*_ = corr(*x*_·*e*_, *y*). This results into a set of edge-wise correlations 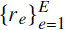 and associated *p*-values computed within the training subjects of that fold. Commonly, edges are then thresholded according to a fixed significance value (e.g., two-tailed *p* < 0.01) to define a set of edges associated with the target score *y*. However, in this study we did not fix a single threshold. Instead, a set of candidate thresholds was proposed, specifically 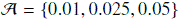, and the feature selection and model-building steps were repeated for each 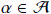.

Then, positive and negative networks were defined by looking at *r*_*e*_. Positive edges are those for which higher connectivity is associated with higher psychopathy scores, whereas negative edges show the opposite pattern. Formally, we define the positive and negative edges as

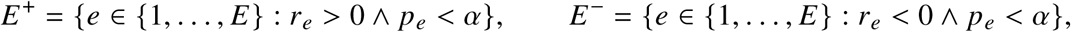

where *α* denotes the chosen threshold. Feature selection is recomputed independently in each cross-validation fold using only the training data so that the information from test subjects does not leak into the model-building stage. The resulting edge sets *E*^+^ and *E*^−^ form the basis for the computation of network strength scores. The selected predictive edges also provide the building blocks for later analysis of psychopathy-related connectivity patterns (H2.1).

To transform the high-dimensional connectivity into low-dimensional predictors, CPM aggregates the selected edges into summary scores, known as “network strength” scores. For each cross-validation split, we compute positive and negative network strengths for every subject *i* by summing the connectivity values of the edges in the corresponding sets *E*^+^ and *E*^−^:

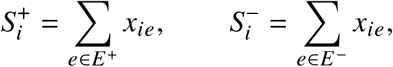

where *x*_*ie*_ denotes the connectivity strength of edge *e* for subject *i*. The resulting scalars 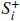 and 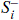 quantify, respectively, the overall strength of edges where higher connectivity is associated with higher psychopathy scores and those where lower connectivity is associated with higher scores. These network strengths are the standard CPM feature summaries and have been shown to provide robust and interpretable predictors that retain information from connectivity patterns while greatly reducing dimensionality.

To test Hypothesis 1.1 that whole-brain rsFC patterns predict Total, Factor 1, and Factor 2 psychopathy scores within the Japanese community sample, a nested cross-validation (CV) was implemented for the CPM. Specifically, *K*_outer_ = 5 outer folds, and within each outer training set, *K*_inner_ = 5 inner folds for parameter selection and edge stability assessment were employed. The outer CV loop intends to provide an unbiased estimate of the out-of-sample predictive performance, while the inner loop is used to select the best feature-selection threshold *α* in 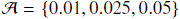 and to quantify the stability of predictive edges. Notably, to provide a better validated metrics, the outer CV was repeated 50 times.

For each outer training set the CPM was performed within the inner folds as follows:

1. In each inner training fold, feature selection was executed for each candidate threshold 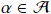, obtaining edge sets *E*^+^ (*α*) and *E*^−^ (*α*), and then the corresponding network strengths *S*^+^ (*α*) and *S*^−^ (*α*) were computed.
2. For each alpha, a linear regression model was fitted on the inner fold train set

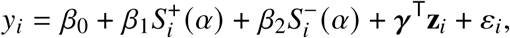

where *y*_*i*_ denotes the psychopathy score of subject *i*, **z**_*i*_ - the covariates (when included), *β*_0_ the intercept, *β*_1_ and *β*_2_ the coefficients for the positive and negative CPM networks, and ***γ*** the coefficients for covariates.

3. Using the fitted coefficients, predictions were generated in the inner validation folds, and the Spearman correlation was computed between predicted and observed scores for each alpha:

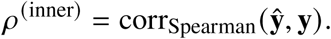

The threshold *α*^∗^ that yielded the best average inner-loop performance was selected for that outer training set:

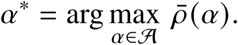

4. In parallel, for each edge the algorithm recorded how often it was selected into the positive and negative networks across the inner folds with the optimal threshold *α*^∗^.

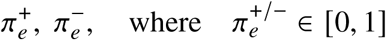

is the proportion of inner folds in which edge *e* appeared in *E*^+/−^ (*α*^∗^).

5. Then, the stability threshold of 0.6 was applied to define the final sets of stable positive and negative edges:

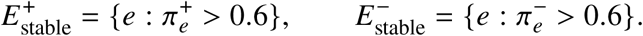

In this way, only edges that were consistently selected across inner folds were carried forward to the outer loop.

For each outer fold, where the test subjects were held out completely, the CPM model was fitted on the entire outer training set using the stable edge sets derived from the corresponding inner loop:

1. Using 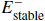 and 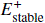, the algorithm computed positive and negative network strengths for all subjects in the outer training and test sets 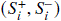.
2. Next, the CPM regression model was fitted on the outer training subjects

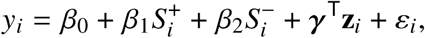

Where *y*_*i*_ denotes the psychopathy score of subject *i*, **z**_*i*_ the covariates (if included), *β*_0_ the intercept, *β*_1_ and *β*_2_ the coefficients for the positive and negative CPM networks, ***γ*** the covariate coefficients, and *ε*_*i*_ the error term.

3. In addition to the model outlined above, separate linear regression models were used for negative or positive network summary scores, thus providing separate linear models for each network type.
4. To investigate the effect of covariates, a model using only covariates to predict scores was implemented.
5. The fitted coefficients (*β*_0_, *β*_1_, *β*_2_, ***γ***) were then applied to the network strengths of the outer test subjects to obtain out-of-sample predictions 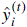.

Additionally, a confound-corrected[63] variant of the model was implemented in which the effects of covariates were regressed out of the CPM network-strength features before prediction. For each network (positive, negative, both) and each subject *i*, *C*_*i*_ denotes the corresponding network-strength vector, and **z**_*i*_ the covariate vector. Within the outer training set, a linear regression model was fitted:

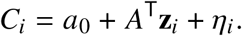

It resulted in covariate-adjusted CPM features as the residuals:

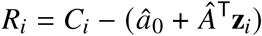

where *â*_0_ and *Â* are the regression coefficients estimated from the outer training data. The same fitted model was then used to compute residualized features *R*_*i*_ for the outer test subjects.

Using these predictors, four related linear models were estimated in each outer fold: (1) a connectome-only model using *C*_*i*_ as predictors; (2) a covariates-only model using *z*_*i*_; (3) a residuals-only model using the confound-regressed features *R*_*i*_; and (4) a full model that combined CPM features and covariates. Comparing performance across these models allowed us to quantify how much variance in psychopathy scores was explained by covariates alone, by CPM features alone, and by CPM features after removal of linear covariate effects, while still basing all performance estimates on outer-fold predictions for unseen subjects.

To test Hypothesis 3.1, the CPM model was trained on the Japanese community sample. The German cohort was then used as an external out-of-sample test set to examine whether the connectivity-psychopathy associations learned in the Japanese sample generalized to a different cultural context.

#### 3.2.2 Weighted CPM (exploratory)

This subsection addresses research question 4, namely whether a weighted-sum implementation of CPM improves prediction relative to the standard unweighted network-strength model. As established earlier, in the baseline CPM, all edges within the positive and negative networks contribute equally to the network-strength scores (sum of selected edges). This implicitly assumes that edges with weak and strong brain-behavior associations are equally informative. However, in weighted CPM, we instead assign larger weights to edges whose connectivity shows stronger associations with the psychopathy scores in the training data. In this way, the network-strength scores are more heavily driven by edges that consistently covary with the outcome, while edges with weaker or more unstable associations contribute proportionally less.

For each fold, network (positive/negative) and target score, the algorithm took the correlation coefficient *r*_*e*_ between edge *e* and the behavioral score (estimated in training data, see Section 3.2.1) and computed their absolute values |*r*_*e*_ | within the set of selected edges. Edges were then ranked in descending order of |*r*_*e*_ | with ranks *R*_*e*_ = 1, 2, 3, The weight was defined as an inverse of this rank: 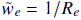 which were normalized within each network so that they sum to 1.

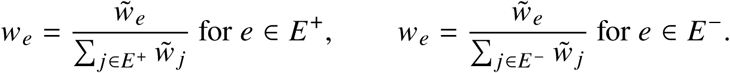

This allowed for the largest |*r*_*e*_ | edge to receive the largest weight. Weighted positive and negative network strengths for subject *i* were then computed as

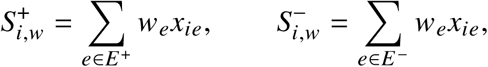

where *x*_*ie*_ is the connectivity value of edge *e* for subject *i*. These weighted scores replace the unweighted sums used in the baseline CPM but are otherwise entered into the same linear regression framework with nested cross-validation described in Section 3.2.1.

#### 3.2.3 Network-Based Statistic+CPM (exploratory)

This subsection addresses Research Question 5, specifically whether subnetworks derived with a network-based statistic (NBS) approach provide different or improved predictions compared with conventional CPM edge selection. The NBS was originally proposed as a way to detect clusters of connected edges that are associated with a variable of interest, while controlling family-wise error at the component level rather than at individual edges[37]. In standard NBS, one first applies a primary edge-wise threshold to a connectivity statistic, then identifies connected components (subnetworks) of suprathreshold edges, and finally uses permutation testing to assess whether the size of each component is unlikely under null. This makes NBS particularly suited to finding spatially extended topologically coherent connectivity patterns. Recent developments in the field of connectome-based machine learning introduced an NBS-Predict algorithm that extends the idea by combining NBS components with machine learning models which are used to predict behavior. This approach is thought to improve performance and interpretability, since features come as connected subnetworks that explicitly include network topology information into the modelling process[38].

The proposed NBS+CPM approach follows the same framework as baseline CPM, but instead of univariate feature selection, it restricts the selected edges to those that belong to coherent subnetworks. The rest of the CPM pipeline (network-strength computation, linear models, nested cross-validation) is left unchanged.

Within each training set of the nested cross-validation (inner and outer folds), NBS+CPM defines positive and negative subnetworks in the following steps:

1. For each edge *e*, a Pearson correlation *r*_*e*_ between connectivity and scores was computed and transformed into a t-statistic

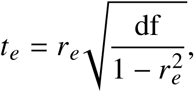

with df = *N* − 2, where *N* is the number of subjects in that training set.

2. Next, a two-tales primary threshold *α* = 0.05 was applied to these statistics, keeping edges with |*t*_*e*_ | > *t*_crit_ corresponding to *p* < 0.05. Suprathreshold edges with *t*_*e*_ > 0 form the positive graph, and those with *t*_*e*_ < 0 form the negative graph, where parcels represent nodes and suprathreshold connections as edges.
3. The suprathreshold graphs obtained can contain many small “whiskers” (single edges or chains of 2-3 edges) that are weakly embedded in the overall network. To focus on more cohesive subnetworks, two graph-theoretic operations were applied:

(a) K-score pruning, which restricts each graph to its k-core with *k* = 2. A k-core is the largest subgraph in which every node has degree at least *k*. Operationally, nodes with fewer than two suprathreshold connections are iteratively removed until all remaining nodes satisfy deg(*v*) ≥ 2 for all *v*
(b) Bridge pruning, which removes edges that act like bridges - edges whose removal would disconnect the graph into two components. Therefore, all such bridge edges were removed using a standard depth-first search (DFS) algorithm. The resulting subnetworks come out as more internally cohesive and less dependent on single “critical” links.
4. Lastly, the connected components were identified using DFS applied to the pruned graph. For each positive and negative sign, the largest connected component by edge count was selected (denoted 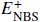 and 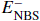). The others were discarded.

Across inner folds, we sought subnetworks that are reproducible rather than constrained to a single split. For each psychopathic score and sign (*s* ∈ {+, −}), we therefore performed component-aware stability based on overlaps of edge sets across inner folds. Let *F* be the number of inner folds. For each fold *f* = 1,…, *F* and sign *s*, denote by 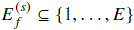 the edge set of the NBS subnetwork obtained in that fold, where *E* is the total number of possible edges. Similarity between subnetworks from folds *f* and *g* was quantified using Jaccard index

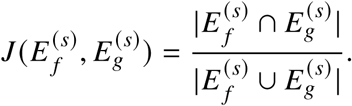

In the current process, two fold-specific subnetworks were considered to belong to the same component if 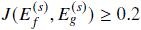. At least 60% of folds have to form a set of clusters with Jaccard similarity ≥ 0.2. Next, edge-wise stability across inner folds was computed. Edges were retained if their stability index is ≥ 0.6.

After enforcing that this consensus again forms a single largest connected component, we obtain stable NBS subnetworks 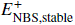, 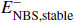. In each outer training set, these subnetworks are summarized into network strength scores for subject *i*,

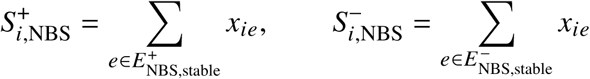

and these replace the baseline CPM strengths in the same family of connectome-only, residuals-only, and full linear models implemented. Prediction and nested cross-validation proceed exactly as described in Section 3.2.1. To address research question 5, we then compare NBS+CPM and standard CPM, examining the models’ predictive performance.

### 3.3 Evaluation metrics

This section defines how we quantify “successful prediction”, how we test Hypothesis 1.1 (prediction within the Japanese sample) and Hypothesis 3.1 (generalization in the German sample), and how we interpret exploratory model variants (weighted CPM and NBS+CPM; research questions 4-5).

For each model, psychopathy score and dataset, predictive performance was evaluated using out-of-sample predictions from the outer folds of the nested cross-validation. Let *y*_*i*_ denote the observed score for subject *i* and *ŷ*_*i*_ the corresponding predicted score. Across all subjects per fold, the metrics computed were:

1. Pearson correlation between predicted and observed scores,

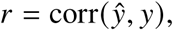

which is the conventional primary performance metric in CPM.

2. Mean absolute error (MAE)

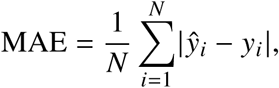

which quantifies the absolute deviation of predictions from the observed scores in the target variable’s original units.

3. Coefficient of determination (*R*^2^)

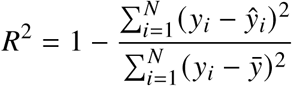

representing goodness-of-fit measure for linear regression models, where *ŷ* is the mean observed score across subjects.

All metrics were computed on test data only; no subject contributed both to the training and test sets for a given outer-fold prediction. When models were repeated multiple times, their performance was summarized as the mean and standard deviation of the metrics.

#### 3.3.1 Permutation testing

To test whether predictive performance exceeded what would be expected by chance, permutation testing was employed at the level of the CPM pipeline. For each psychopathy score and model configuration, a null distribution of performance was constructed by repeatedly shuffling the correspondence between functional connectivity and outcome scores and rerunning the full analysis:

1. For a given dataset and outcome variable, *B* = 5000 permutations were generated. In permutation *b*, the psychopathic scores across subjects were randomly permuted, obtaining a vector *y*^(*b*)^ that preserves the marginal distribution of scores, but removes any true association with the connectomes.
2. For each permuted outcome *y*^(*b*)^, the entire CPM pipeline was rerun, including nested cross-validation, feature selection, network strength computation, model fitting, and out-of-sample prediction, exactly as for the original data. This yielded permuted performance metrics for each permutation, such as correlation scores or error metrics, which we denote *m*^(*b*)^.

The collection of permutation metrics defines the null distribution of predictive performance under the hypothesis that rsFC and psychopathic scores are unrelated. For the observed (non-permuted) data, the actual performance is computed.

3. To assess the significance of the models’ performance, for each metric and model, the algorithm compared the observed performance *m*_obs_ (from the non-permuted data) to the set of permuted values 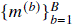.

(a) For score metrics, where higher values indicate better prediction, the permutation p-value was defined as

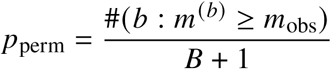

indicating a fraction of permutation runs in which a model trained on permuted outcomes achieved a higher score than the model trained on the true outcomes.

(b) For error metrics, where lower values indicate better prediction, the inequality was reversed:

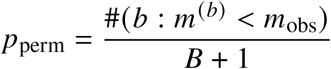

corresponding to the fraction of permutation runs in which a model trained on permuted outcomes achieved a smaller error than observed.

In this way, *p*_perm_ quantifies how often a model trained on shuffled labels outperformed the model trained on true labels. Hypothesis 1.1 (prediction within the Japanese sample) and Hypothesis 3.1 (prediction within the German sample) were considered supported only when permutation p-values for the baseline model were below the significance threshold (*p* < 0.05).

In addition, for the stability analyses, null distributions were also constructed for edge stability across permutations. For each edge, the proportion of permutation runs was computed in which its stability was at least as high as in the non-permuted data, yielding an edge-wise stability p-value.

## 4 Results

To test the central hypotheses regarding the neural underpinnings of psychopathy, we first implemented a connectome-based predictive modeling approach. More precisely, we examined whether resting-state functional connectivity could predict the total psychopathic score, as well as Factor 1 and Factor 2, in a Japanese community sample. Second, in a set of exploratory analyses, we evaluated (a) alternative model implementations (weighted CPM and NBS + CPM), (b) potential sex differences, and (c) cross-dataset generalization to an independent German cohort. Finally, we also fitted CPM models for the four SRP-SF facet scores to explore more fine-grained relationships between functional connectivity and psychopathy traits.

### 4.1 RQ1: Prediction of Psychopathic Traits from rsFC (H1.1.)

During model training, CPM showed an excellent fit to the data. Across cross-validation folds, the average Pearson correlation, which quantifies the linear association between predicted and observed values, in the training sets was high (mean *r* ≈ 0.85 for Total, Factor 1 and Factor 2).

However, contrary to my main hypothesis, this fit did not generalize to the test data. CPM models trained on whole-brain functional connectivity did not yield significant out-of-sample predictions for total, Factor 1, or Factor 2 scores. As detailed in Table 1, the average test-set Pearson’s *r* across all cross-validation folds was near zero.

**Table 1:**
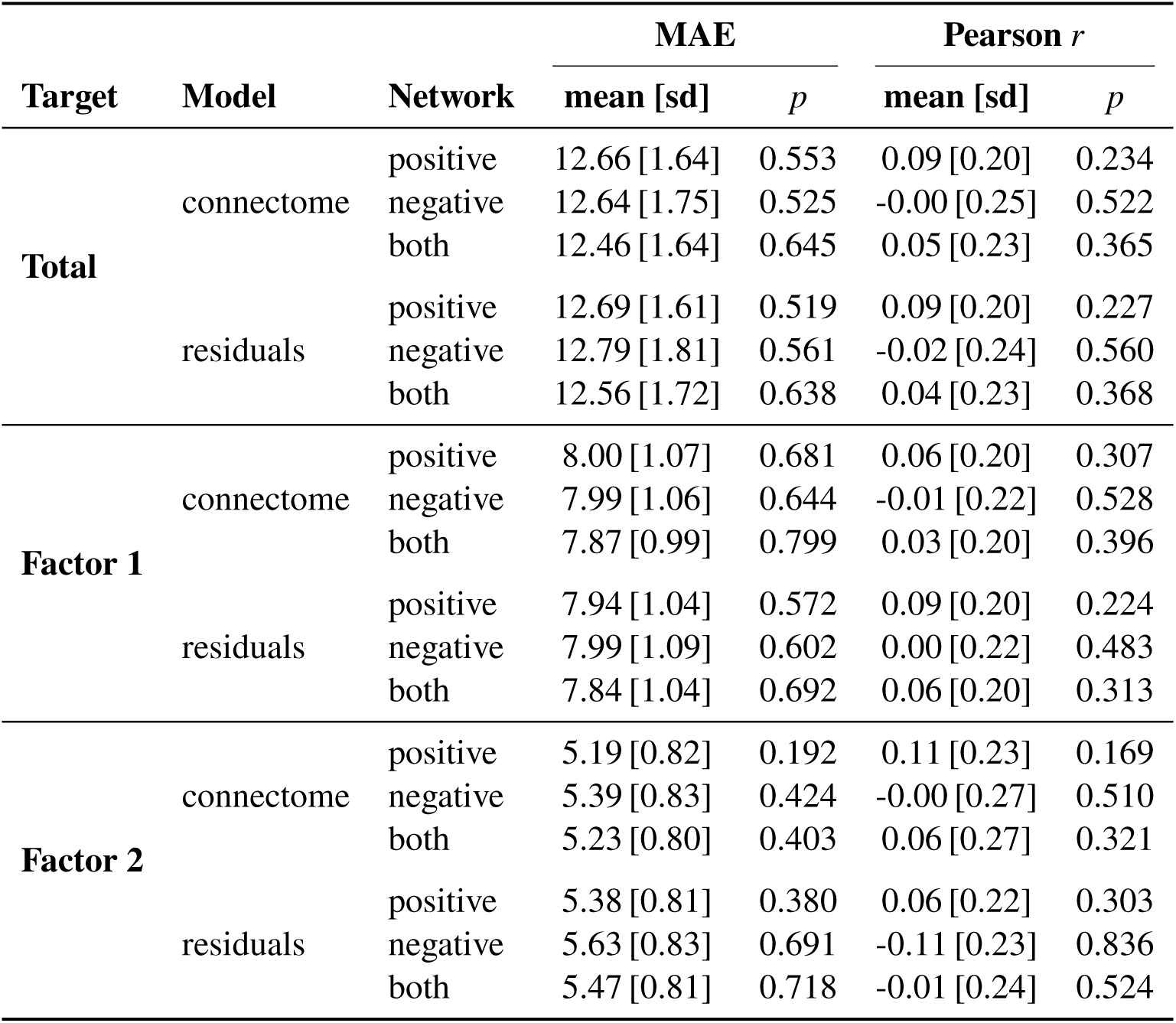
Baseline CPM performance (mean [sd] across CV repeats) with permutation *p*-values.

Permutation tests supported the null result by generating a null distribution of test-set correlations and calculating the proportion of these null correlations that are at least as large as the one observed with the true data. All permutation p-values were well above the significance threshold ((*p* > 0.2)), meaning that the observed performance is not different from the one generated by chance. Furthermore, the mean absolute error (MAE) values were comparable to a mean-prediction baseline (e.g., Total score baseline ≈ 11.4; Factor 1 ≈ 7.0; Factor 2 ≈ 4.9). This shows that models did not outperform trivial predictions of the sample mean.

Figure 3 illustrates a representative cross-validation repeat, comprising subsequent 5 folds in the cross-validation procedure, where test predictions cluster around the sample mean, consistent with negligible predictive variance and a lack of linear relationship.

**Figure 3:**
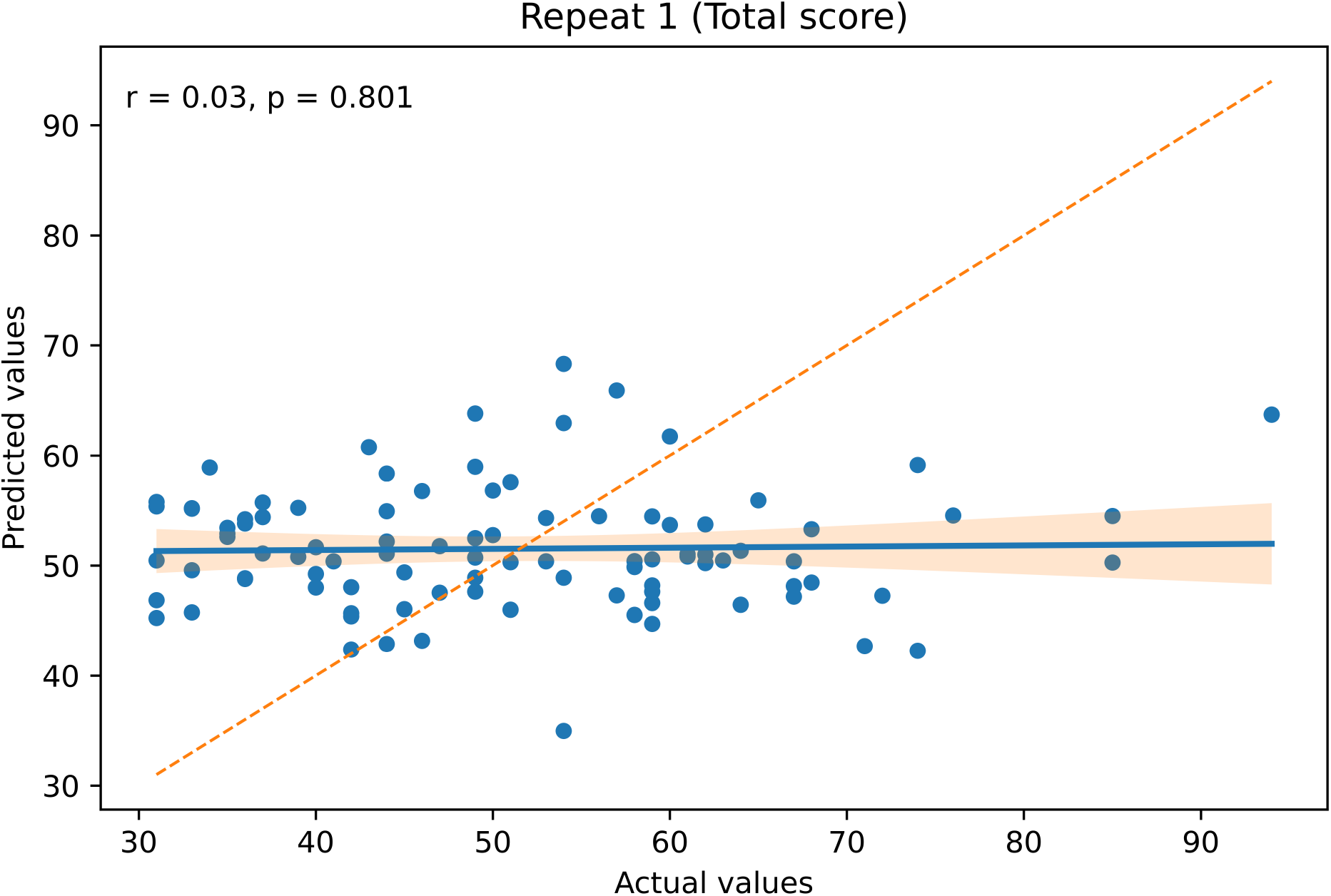
Example test set scatter plot from a single cross-validation repeat, showing the relationship between actual and predicted Total scores.

### 4.2 RQ2: Contributing Resting-state Networks (H2.1.)

I originally sought to examine whether any predictive effects would be driven by connections within particular intrinsic resting-state networks (default mode, salience, frontoparietal control). However, this analysis was contingent on a significant finding for RQ1. Since the target baseline CPM models for psychopathy Total and Factor scores did not achieve significant out-of-sample performance (see Section 4.1), Hypothesis 2.1 could not be tested and no network-level conclusions are drawn.

### 4.3 RQ3: Generalization to an Independent Cohort (H3.1.)

I also sought to test whether a CPM model trained on a Japanese community sample would generalize to German participants from an independent community cohort. This required a reliable predictive model developed using the Japanese data that could then be applied, unchanged, to the German individuals. As reported in Section 4.1, however, the models showed no significant out-of-sample prediction during cross-validation. Therefore, Hypothesis 3.1 could not be tested as originally specified.

However, to see whether the null findings persisted across cohorts, we next ran the full CPM pipeline independently on the German dataset, using SD3 psychopathy subscale scores as the target variable for prediction. The results were similarly null, with cross-validated correlations between predicted and observed scores being near zero, permutation tests being non-significant, and mean absolute errors being comparable to a mean-prediction baseline (see Table 2). Null findings persisted even when restricting the age range to match the Japanese sample or when stratifying by sex, meaning resting-state functional connectivity did not significantly predict the severity of psychopathic traits.

**Table 2:** German dataset — MAE and Pearson *r* (mean [sd] across CV repeats) with permutation *p*-values for *connectome* and *residuals*.

| Target | Model | Network | MAE | | Pearson $r$ | |
| --- | --- | --- | --- | --- | --- | --- |
| | | | mean [sd] | $p$ | mean [sd] | $p$ |
| All data | connectome | positive | 3.77 [0.54] | 0.157 | 0.13 [0.21] | 0.128 |
|  |  | negative | 3.83 [0.56] | 0.238 | 0.01 [0.22] | 0.473 |
|  |  | both | 3.66 [0.53] | 0.103 | 0.11 [0.22] | 0.178 |
|  | residuals | positive | 4.19 [0.64] | 0.740 | −0.09 [0.22] | 0.796 |
|  |  | negative | 4.27 [0.61] | 0.848 | −0.17 [0.21] | 0.936 |
|  |  | both | 4.32 [0.61] | 0.937 | −0.17 [0.20] | 0.927 |
| Youth only | connectome | positive | 3.85 [0.59] | 0.307 | 0.05 [0.22] | 0.343 |
|  |  | negative | 3.76 [0.62] | 0.183 | 0.09 [0.22] | 0.237 |
|  |  | both | 3.66 [0.62] | 0.171 | 0.10 [0.22] | 0.236 |
|  | residuals | positive | 4.06 [0.60] | 0.561 | −0.03 [0.20] | 0.562 |
|  |  | negative | 4.06 [0.65] | 0.557 | −0.04 [0.22] | 0.628 |
|  |  | both | 4.04 [0.63] | 0.732 | −0.05 [0.19] | 0.641 |
| Male | connectome | positive | 3.76 [0.74] | 0.094 | 0.14 [0.32] | 0.166 |
|  |  | negative | 4.52 [0.86] | 0.936 | −0.22 [0.26] | 0.941 |
|  |  | both | 3.98 [0.76] | 0.562 | −0.01 [0.29] | 0.534 |
|  | residuals | positive | 4.01 [0.85] | 0.180 | 0.05 [0.34] | 0.365 |
|  |  | negative | 4.62 [0.94] | 0.860 | −0.21 [0.27] | 0.931 |
|  |  | both | 4.22 [0.88] | 0.578 | −0.09 [0.31] | 0.726 |
| Female | connectome | positive | 3.47 [0.77] | 0.777 | −0.11 [0.34] | 0.721 |
|  |  | negative | 3.16 [0.72] | 0.325 | 0.04 [0.36] | 0.410 |
|  |  | both | 3.20 [0.73] | 0.569 | −0.07 [0.35] | 0.626 |
|  | residuals | positive | 3.71 [0.82] | 0.762 | −0.13 [0.36] | 0.757 |
|  |  | negative | 3.41 [0.72] | 0.417 | 0.02 [0.33] | 0.451 |
|  |  | both | 3.43 [0.76] | 0.594 | −0.08 [0.34] | 0.668 |

### 4.4 RQ4 & RQ5: Alternative Model Implementations

I tested whether alternative modeling approaches, such as weighted-edge CPM and NBS+CPM, could improve performance over the baseline model. Neither implementation improved predictive accuracy relative to the baseline model. Test set Pearson’s r values remained near zero for total and factor scores (see Table 3).

**Table 3:** Comparison of CPM variants using the combined edge set (“both”) for connectome and residuals models (mean [sd] across CV repeats) with permutation *p*-values.

| Target | Variant | Connectome |  |  |  | Residuals |  |  |  |
| --- | --- | --- | --- | --- | --- | --- | --- | --- | --- |
| | | MAE | | Pearson $r$ | | MAE | | Pearson $r$ | |
| | | mean [sd] | $p$ | mean [sd] | $p$ | mean [sd] | $p$ | mean [sd] | $p$ |
| **Total** | Baseline CPM | 12.46 [1.64] | 0.645 | 0.05 [0.23] | 0.365 | 12.56 [1.72] | 0.638 | 0.04 [0.23] | 0.368 |
|  | Weighted-edge CPM | 12.95 [1.69] | 0.791 | −0.04 [0.21] | 0.633 | 13.08 [1.78] | 0.808 | −0.06 [0.22] | 0.720 |
|  | NBS+CPM | 12.31 [1.68] | 0.593 | 0.08 [0.24] | 0.277 | 12.34 [1.70] | 0.601 | 0.08 [0.23] | 0.283 |
| **Factor 1** | Baseline CPM | 7.87 [0.99] | 0.799 | 0.03 [0.20] | 0.396 | 7.84 [1.04] | 0.692 | 0.06 [0.20] | 0.333 |
|  | Weighted-edge CPM | 8.14 [1.08] | 0.866 | −0.02 [0.20] | 0.576 | 8.10 [1.12] | 0.784 | −0.01 [0.21] | 0.535 |
|  | NBS+CPM | 7.76 [1.00] | 0.754 | 0.06 [0.22] | 0.311 | 7.79 [1.01] | 0.765 | 0.02 [0.21] | 0.445 |
| **Factor 2** | Baseline CPM | 5.23 [0.80] | 0.403 | 0.06 [0.27] | 0.321 | 5.47 [0.81] | 0.718 | −0.01 [0.24] | 0.524 |
|  | Weighted-edge CPM | 5.32 [0.81] | 0.388 | 0.03 [0.24] | 0.385 | 5.54 [0.83] | 0.671 | −0.03 [0.24] | 0.726 |
|  | NBS+CPM | 5.18 [0.77] | 0.367 | 0.08 [0.27] | 0.243 | 5.17 [0.79] | 0.339 | 0.09 [0.27] | 0.235 |

### 4.5 RQ6: Sex Differences

To determine whether sex moderated predictive accuracy, we tested for sex differences in the primary models. we stratified the model by sex to test whether rsFC predicted total, Factor 1, or Factor 2 scores across males and females separately. The developed models did not yield significant results for either sex.

### 4.6 Post Hoc Analyses

The consistent null findings in my planned analyses (RQ1 - RQ6) motivated a set of further, unhypothesized analyses to understand potential reasons behind the null findings. we explored (a) whether a predictive signal existed at a more granular facet level and (b) whether the null results could be explained by data heterogeneity.

#### 4.6.1 Antisocial Facet Predictive Signal

Given that CPM yielded null results for the total, Factor 1, and Factor 2 scores, we applied CPM to the four facets - interpersonal manipulation, callous effect, antisocial behavior, and erratic lifestyle - to investigate whether fine-grained dimensions would yield more precise findings. Among facet CPM models, only the antisocial behavior facet showed significant predictions on average across folds (see Table 4). The model based on positive network - edges whose connectivity strength was positively correlated with antisocial scores - showed modest but reliable performance (*r* = 0.27[0.24], *p* = 0.007**, MAE = 1.94[0.41]). A combined positive-plus-negative network model performed similarly (*r* = 0.27[0.31], *p* = 0.012*, MAE = 1.94[0.4]). In contrast, the negative network did not pass the permutation testing (*r* = 0.16[0.32], *p* = 0.073, MAE = 2.13[0.38]). This effect was weaker but remained significant even when predicting the variance in antisocial behavior after regressing out covariates also produced significant but weaker correlations (pos.n.: *r* = 0.2[0.24], *p* = 0.038*, MAE = 1.94[0.41]; neg.n.: *r* = 0.14[0.32], *p* = 0.111, MAE = 2.18[0.38]; pos.n + neg.n.: *r* = 0.2[0.3], *p* = 0.045*, MAE = 2.04[0.4]). Furthermore, it was found that the alternative models (weighted CPM and NBS+CPM) offered no clear improvement for this one significant model. Cross-validated *R*^2^ estimates were, on average, negative, reflecting that the predictions are close to a mean-prediction baseline. The details can be found in Table 4.

**Table 4:** CPM variants predicting antisocial behaviour facet: MAE, Pearson *r*, and *R*^2^ (mean [sd] across CV repeats) with permutation *p*-values. Bold values indicate *p* < .05 (* *p* < .05, ** *p* < .01).

| Model | Modality | Network | MAE | | Pearson $r$ | | $R^2$ | |
| --- | --- | --- | --- | --- | --- | --- | --- | --- |
| | | | mean [sd] | $p$ | mean [sd] | $p$ | mean [sd] | $p$ |
| CPM | connectome | positive | **1.94 [0.41]** | **0.005\*\*** | **0.27 [0.24]** | **0.007\*\*** | **-0.23 [0.53]** | **0.012\*** |
|  |  | negative | 2.13 [0.38] | 0.109 | 0.16 [0.32] | 0.073 | **-0.31 [0.58]** | **0.031\*** |
|  |  | both | **1.94 [0.40]** | **0.010\*\*** | **0.27 [0.31]** | **0.012\*** | **-0.15 [0.49]** | **0.009\*\*** |
|  | residuals | positive | 2.08 [0.40] | 0.071 | **0.20 [0.24]** | **0.038\*** | -0.35 [0.60] | 0.056 |
|  |  | negative | 2.18 [0.38] | 0.189 | 0.14 [0.32] | 0.111 | -0.40 [0.67] | 0.094 |
|  |  | both | 2.04 [0.40] | 0.072 | **0.20 [0.30]** | **0.045\*** | -0.28 [0.61] | 0.085 |
| Weighted CPM | connectome | positive | **2.00 [0.43]** | **0.004\*\*** | **0.26 [0.26]** | **0.008\*\*** | **-0.26 [0.57]** | **0.006\*\*** |
|  |  | negative | 2.13 [0.41] | 0.052 | 0.16 [0.27] | 0.057 | -0.39 [0.65] | 0.051 |
|  |  | both | **1.98 [0.42]** | **0.007\*\*** | **0.25 [0.28]** | **0.008\*\*** | **-0.21 [0.54]** | **0.008\*\*** |
|  | residuals | positive | **2.08 [0.44]** | **0.022\*** | **0.20 [0.26]** | **0.026\*** | **-0.35 [0.63]** | **0.023\*** |
|  |  | negative | 2.16 [0.42] | 0.065 | 0.16 [0.28] | 0.062 | -0.44 [0.72] | 0.069 |
|  |  | both | **2.06 [0.43]** | **0.030\*** | **0.21 [0.28]** | **0.024\*** | **-0.32 [0.63]** | **0.035\*** |
| NBS+CPM | connectome | positive | **1.96 [0.41]** | **0.007\*\*** | **0.27 [0.24]** | **0.009\*\*** | **-0.25 [0.56]** | **0.016\*** |
|  |  | negative | 2.18 [0.37] | 0.252 | 0.14 [0.32] | 0.109 | -0.35 [0.60] | 0.071 |
|  |  | both | **1.97 [0.40]** | **0.024\*** | **0.27 [0.31]** | **0.010\*\*** | **-0.16 [0.50]** | **0.017\*** |
|  | residuals | positive | **2.03 [0.39]** | **0.035\*** | **0.23 [0.25]** | **0.020\*** | **-0.29 [0.56]** | **0.028\*** |
|  |  | negative | 2.18 [0.37] | 0.213 | 0.15 [0.33] | 0.092 | -0.35 [0.62] | 0.067 |
|  |  | both | 2.00 [0.39] | 0.052 | **0.25 [0.31]** | **0.020\*** | **-0.19 [0.52]** | **0.033\*** |

#### 4.6.2 Networks Contributing to the Antisocial Facet Prediction

To identify which networks contributed the most to the antisocial behavior prediction, first, the edges that appeared in at least 80% of the 250 cross-validation folds were selected. From these stable predictive edges, we then computed node degree (hubness) for each ROI, defined as the number of stable predictive edges connected to each region, to quantify its contribution to the model. Regions with a higher degree therefore occurred in more of the most stable predictive connections. As shown in Figure 4, the positive network was dominated by hubs in the default mode network, followed by the control and salience networks. In contrast, the negative network showed the most prominent hubs in the control and default mode networks. Overall, the hub analysis revealed that both predictive networks, despite their opposite relationships to the antisocial score, converged on the same core systems: the default mode and frontoparietal control networks.

**Figure 4:**
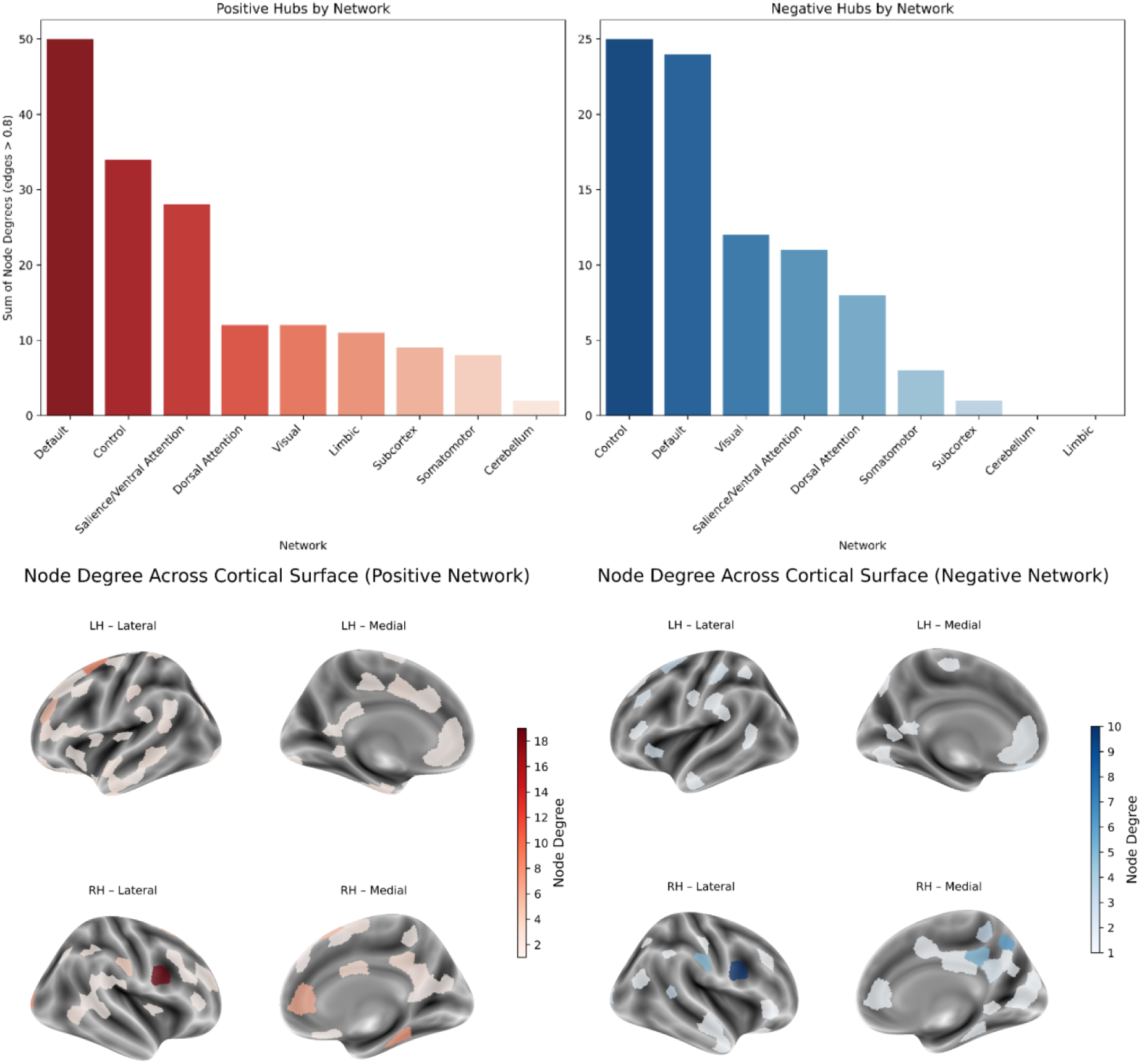
Networks Contributing to the Antisocial Facet Prediction. (a) Network-level and (b), (c) spatial distribution of stable predictive hubs for the antisocial behavior model.

#### 4.6.3 Investigating Participants’ Heterogeneity (IS-RSA)

Together, the results above suggest that the failure to predict psychopathy scores was not a simple artefact of the baseline pipeline, a peculiarity of the Japanese sample, or a product of unmodeled sex differences. These considerations motivated us to examine one of the possible explanations in more detail: heterogeneity in the relationship between resting-state connectivity patterns and psychopathic traits across individuals.

To investigate whether the poor predictive performance of the CPM models corresponds to a high heterogeneity between brain connectivity and psychopathic traits, we employed inter-subject representational similarity analyses (IS-RSA).

##### Objectives

Recent work in resting-state fMRI has shown how meaningful individual differences can emerge in the pattern of similarity between participants. Particularly, inter-subject representational similarity analysis (IS-RSA) relates subject-by-subject neural similarity to subject-by-subject similarity in behavioral or trait measures[64], and has been used to uncover structure in affective experience[65], temperament[66], grit[67], and other personality traits[68, 69]. IS-RSA is therefore well-suited for asking whether the psychopathy continuum is reflected in the geometry of inter-individual connectivity patterns.

In post-hoc analyses, IS-RSA models were used to address the following questions:

- Do individuals with similar psychopathy scores show more similar functional connectivity patterns (Nearest-Neighbour (NN) model)?
- Do we observe an Anna Karenina (AnnaK) organization, where the brain connectivity of certain regions is especially similar at one extreme of the psychopathy continuum and increasingly diverges away from that extreme?
- Does using item-wise psychopathy profiles (rather than summary scores) strengthen brain-trait coupling, as suggested by recent IS-RSA work[65] using item-level or high-dimensional behavioral representations?

All IS-RSA analyses were conducted at the parcel level. By testing how neural similarity matrices in the defined regions track different behavioral similarity models, these post-hoc analyses provided a follow-up to Hypothesis 2.1, and helped us to interpret why CPM prediction may have failed despite potential structure in the underlying connectivity-psychopathy relationship.

##### Nearest-Neighbor

The first IS-RSA model tested whether individuals who are closer in Total psychopathic score also showed more similar functional connectivity patterns, irrespective of their absolute position on the trait continuum. Nearest-neighbor approach[64] treats psychopathy as a dimension along which both behavior and brain organization vary gradually.

For each region of interest, a neural similarity matrix was derived, that captured how similar subjects’ connectivity patterns are within that region. The behavioral similarity using the NN approach was defined as a monotonically decreasing function of absolute rank distance between individuals on the Total score on the SRP-SF scale. To quantify brain-behavior relationship under the NN hypothesis for each region, the neural and behavioral similarity patterns were correlated using a rank-based Spearman correlation. This procedure resulted into a single coefficient per parcel, where positive values indicate that in region *r*, pairs of subjects who are closer in total psychopathy score also tend to have similar FC.

##### Anna K

The second IS-RSA model was Anna Karenina (AnnaK)[64], which tested the hypothesis that neural responses are especially similar at one extreme of the behavioral spectrum (here, psychopathic traits) and become more diverse toward the opposite end (“All high [or low] scorers are alike; each low [or high] scorer is different in their own way”). In contrast to the nearest-neighbor model, which emphasises relative distance along the scale (e.g., a pair of individuals scoring 1 and 10 are just as similar as individuals scoring 91 and 100), the AnnaK model encodes absolute position on the psychopathy continuum. In this thesis, the AnnaK model was used to test whether there are regions or networks in which high-psychopathy individuals show similar connectivity patterns, whereas lower-psychopathy individuals are more heterogeneous. Positive similarity values indicate that in a specific region, subjects towards the trait’s higher extreme are more similar to one another than to individuals closer to the opposite end of the distribution.

**Item-wise (Chen et al. (2020))** The NN and AnnaK models both encode one-dimensional behavioral scores; however, psychopathic traits are measured via multi-item questionnaires that capture a richer profile than any single total score. Recent IS-RSA work has shown that using item-wise response profiles to construct behavioral similarity matrices can substantially increase brain-behavior coupling compared with univariate scores. we therefore adopted Chen et al. (2020)[65] item-wise IS-RSA as a final approach to test whether local FC patterns track fine-grained psychopathy profiles. Positive brain-behavior similarity values indicate that subjects with more similar item-level psychopathy profiles also have more similar FC patterns in region *r*.

#### 4.6.4 Results of IS-RSA

The Nearest Neighbors (NN) approach, testing whether subjects that are behaviorally similar are also nearby in the connectome space, revealed no regions showing significant correspondence between similarity in psychopathy scores and brain connectivity. Parcel-wise rho values were concentrated around zero, suggesting that individuals with similar psychopathy levels do not share highly similar connectivity profiles. Additionally, none of the parcels survived false-discovery-rate (pFDR) correction (pFDR range: 0.93-0.99).

In contrast, the AnnaK approach, which tests the hypothesis that high-scoring individuals are neurally similar to each other, showed weak brain-behaviour similarity scores in regions of the default mode, salience/ventral attention, and frontoparietal control networks (see Figure 5). After FDR correction, parcel-wise pFDR values ranged from 0.65 to 0.99, and no parcel reached significance.

**Figure 5:**
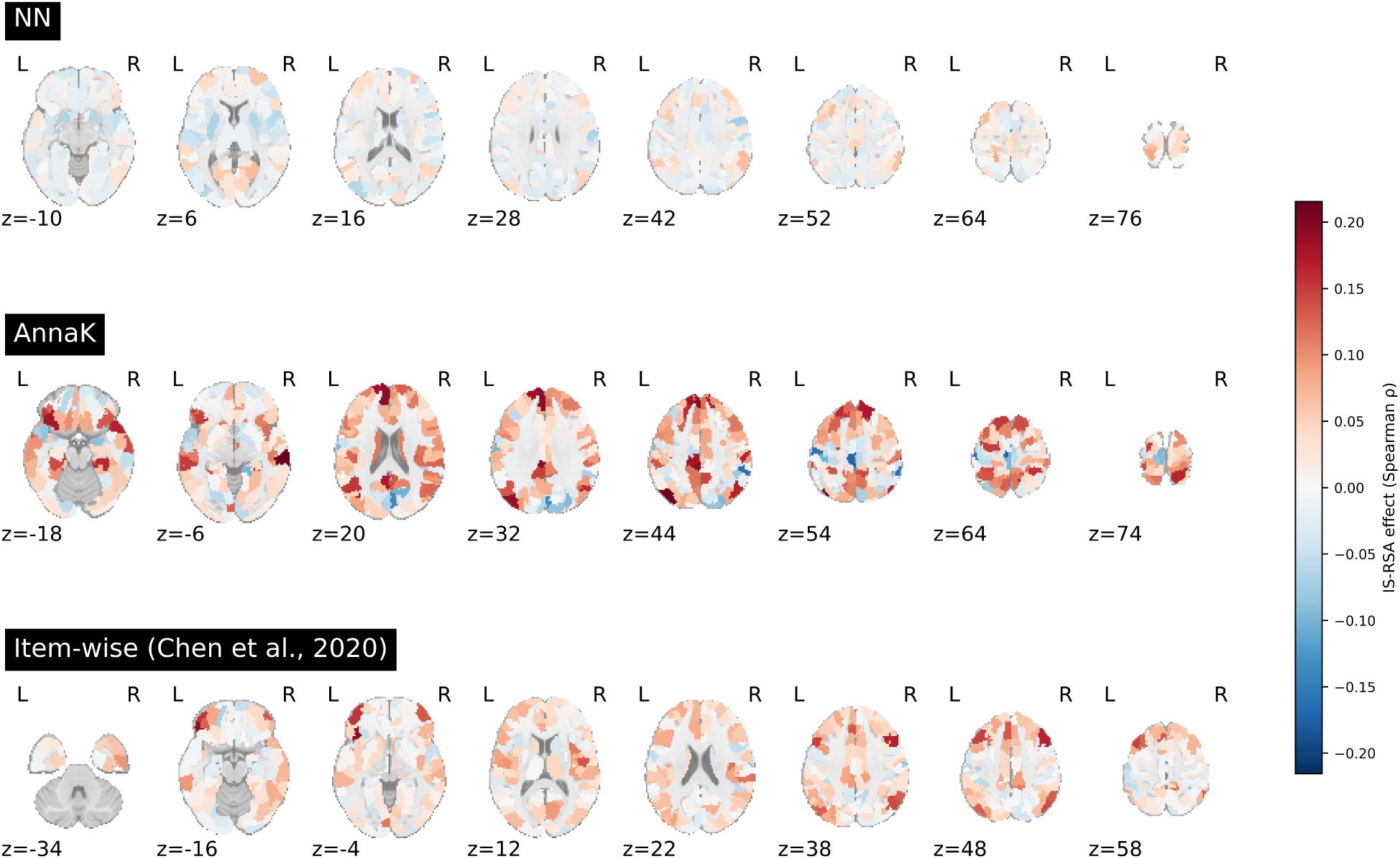
Whole-brain IS-RSA correlation maps for the NN, AnnaK, and item-wise (Chen et al. 2020) methods. Maps depict parcel-wise Spearman rho values representing the correspondence between neural similarity and behavioral similarity (measured by SRP-SF questionnaire) across subjects

Finally, the item-wise approach (Chen et al., 2020), which aligns multivariate questionnaire patterns with neural similarity, suggests weak brain-behavior similarity scores that appear again in DMN, SN, and FPN (see Figure 5). FDR corrected p-values spanned 0.08-0.95, with no parcel surviving the correction.

To summarize the spatial trends, we performed network enrichment analyses of the top parcels exhibiting similar connectivity patterns in similar psychopathy levels (Fig. 6). Parcels belonging to default mode, salience, and frontoparietal control networks were overrepresented among the top-ranked regions, consistent with the patterns in the statistical maps (Fig. 5). Crucially, however, none of the brain regions identified in any IS-RSA approach survived FDR correction, so the results should be interpreted with caution.

**Figure 6:**
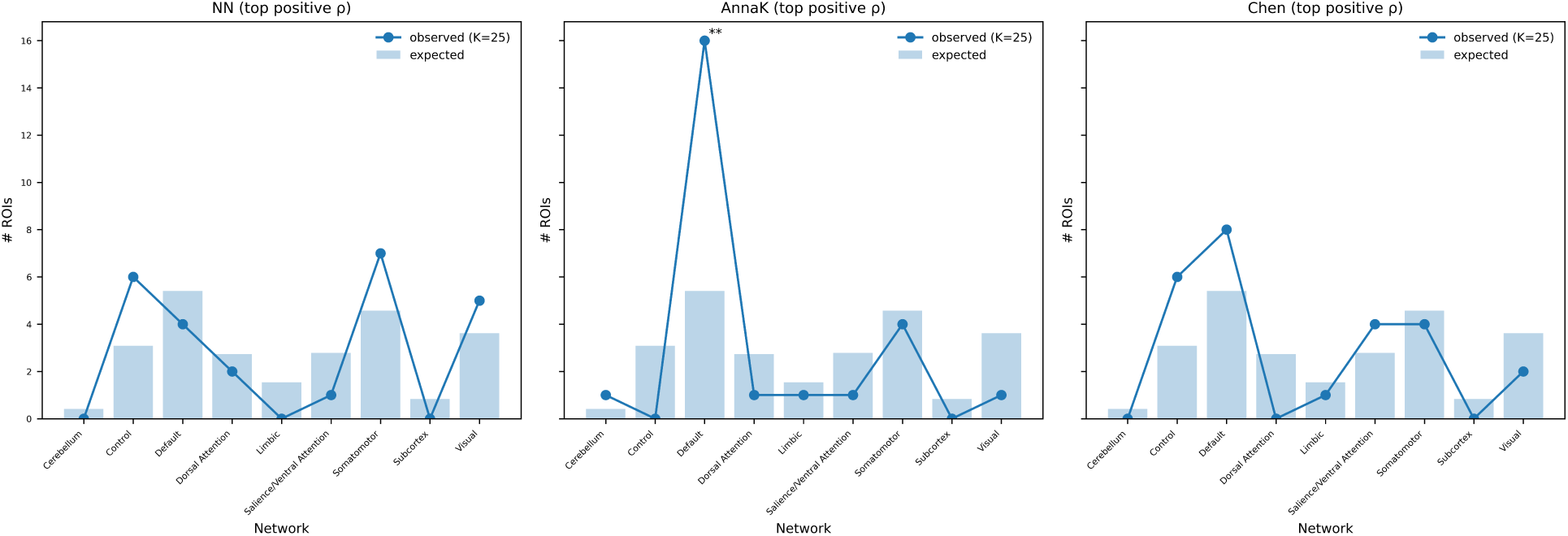
Network enrichment analysis of the top 25 positive IS-RSA regions across NN, AnnaK, and item-wise (Chen et al. 2020) methods. Blue lines represent the number of high-rho ROIs w.r.t. defined functional networks. Shaded bars reflect expected counts based on network size. Asterisks indicate networks with significant over-representation of top-25 parcels (∗*p* < .05, ∗ ∗ *p* < 0.01, uncorrected) (*r* = 0.19, *p* < 0.001).

As a final exploratory step, we investigated whether these spatial patterns aligned with previously reported functional networks. we computed the correlation between the parcel-wise IS-RSA effects and the “Psychopathy network” identified by Dugré and De Brito (2025). As shown in Figure 7, IS-RSA effects were positively, though weakly, correlated with the psychopathy network values

**Figure 7:**
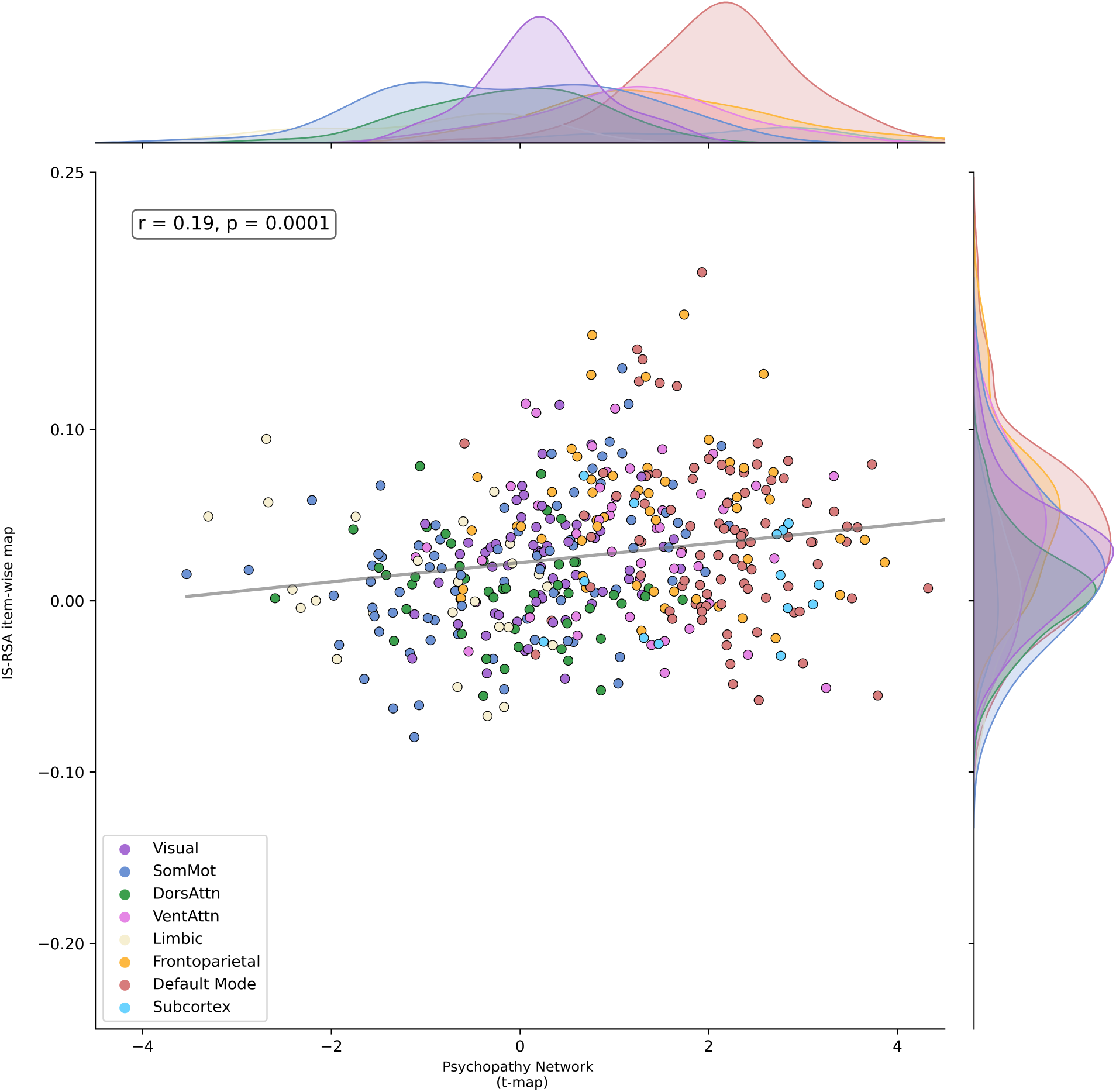
Correlation between parcel-wise IS-RSA (Chen et al., 2020) effects and the psychopathy network (Dugré et al., 2025). Each point represents a brain region colored by its assigned functional network.

## 5 Discussion

### 5.1 Overview of main findings

Over the past decades, neuroimaging studies have shown that individuals high in psychopathic traits often display altered brain structure and function[16, 70, 71]. Yet, most work has relied on relatively small, male-dominated forensic samples from the WEIRD population, leaving open the question of how replicable these brain-behavior associations are in community and non-Western populations. Against the established background, the present thesis investigated whether whole-brain resting-state functional connectivity can predict individual differences in psychopathic traits in community samples. Specifically, the primary aim was to test the predictive ability of functional connectivity in predicting psychopathic traits in a Japanese sample. Another goal was to identify which large-scale networks, if any, carry the predictive signal. Because most neuroimaging research in this field has been conducted in WEIRD samples, the study also explored cross-cultural generalization by testing the model on an independent German sample. As exploratory analyses, we also aimed to look into whether different aggregation or feature selection techniques might improve the model’s performance (weighted CPM and NBS-CPM). Finally, the potential sex differences were investigated by applying the same modelling to females and males separately.

The results from these varied analyses offer several specific insights. Connectome-based predictive modeling did not yield a robust, generalizable predictive model for total and factor scores in a Japanese sample. Since the predictive model could not be evaluated using the German dataset as initially planned, CPM was applied independently on the German dataset to see whether null findings persisted across cohorts. The null results from the primary dataset were also replicated in an external German dataset, indicating that the lack of strong resting-state FC association with the psychopathy traits was not limited to a single cultural group. Moreover, sex differences were not the reason for models’ underperformance, as even after splitting the datasets into males vs females, no significant predictive signal was found. Additionally, neither weighted edge aggregation nor network-based statistics as a feature selection technique impacted the null results of the models. However, a modest effect was found between functional connectivity and the antisocial behavior facet of SRP-SF, where connections in default mode, salience, and frontoparietal networks contributed to the predictive signal.

Due to the consistent null findings throughout the hypothesized analyses, we performed a set of post-hoc analyses to understand the potential reasons behind the null results. Specifically, we utilized inter-subject representational similarity analysis to determine whether the psychopathic traits are reflected in the pattern of similarity between individuals’ connectomes. The IS-RSA revealed weak correspondence between neural and behavioral similarity, concentrated primarily on the default mode, salience, and frontoparietal networks. Taken together, the results suggest that, while prior studies have shown that whole-brain connectivity may be a potential marker for psychopathy in a community sample[30], the relationship is rather complex.

### 5.2 Results in context with previous neuroimaging findings

Task-based and resting-state fMRI studies have linked psychopathy to altered activity and connectivity in a set of regions primarily encompassing default mode, salience, and frontoparietal networks, including the amygdala, hippocampus, ventromedial and dorsomedial prefrontal cortex, posterior cingulate/precuneus, insula, and striatum[14, 20, 70, 72]. In addition, a recent connectome-based predictive modelling study in Chinese college students[30] showed that whole-brain resting-state functional connectivity can predict individual differences in psychopathic traits, with predictive edges centered in anterior prefrontal, orbitofrontal, anterior cingulate, and insular cortices. Specifically, connectivity between the occipital network and the cingulo-opercular network emerged as the strongest predictor of individual differences in psychopathy severity[30].

Despite significant advances in unveiling the neurobiology of psychopathy, there is still no robust biomarker of psychopathy. Critical reviews have already highlighted that structural and functional neuroimaging findings have been disparate, implicating a wide set of regions that are typically obtained from small samples, with diverse tasks and measurements[20, 72]. This brought researchers’ attention to the inconsistent picture of neuroimaging findings in psychopathy. Koenigs et al., for instance, summarized functional MRI work showing that psychopathy is associated with abnormal activity across all four lobes and multiple subcortical regions, noting that “it is difficult to group the findings in any particular functional domain”[72]. Similarly, Pujol et al. synthesized anatomical and functional alterations in the brains of psychopathic individuals and emphasized that there are notable differences in the evidence provided by the existing studies, suggesting a possible biological heterogeneity of psychopathy[20].

More recent systematic reviews and meta-analyses have converged on a similar view. Johanson et al. (2020) reviewed 118 MRI neuroimaging studies (structural, diffusion, and functional) and concluded that although a default mode network consistently recurs across studies, the overall functional pattern is highly heterogeneous, with variability in which regions show associations and in which direction[70]. Deming and Koenigs (2020) task-fMRI meta-analysis identified increased task-related activity in midline cortical regions overlapping the default mode network and medial temporal lobe, alongside decreased activity in dorsal anterior cingulate cortex, which is a key salience network node. At the same time, their findings challenged amygdala hypoactivity and highlighted that consistent abnormalities are limited to a relatively small set of regions along with consistent variability elsewhere[14]. Finally, Deming et al. (2022) directly examined the amygdala findings and concluded that most MRI studies find null relationships between psychopathy and amygdala structure or function[73].

In the context of all the neuroscience works described above, the present findings fit the broader pattern depicted. Across all analyses, connectome-based predictive modelling did not yield stable predictive relationships for any psychopathy total, factor, or facet score. Even for the antisocial facet, which showed the strongest signal, cross-validated predictions were weak and did not outperform a simple mean-score baseline. Together, these findings suggest that resting-state connectivity associations with psychopathy levels in community samples are likely small and heterogeneous.

The null results obtained in this thesis can be interpreted within the context of brain-wide association studies (BWAS). Gratton et al. (2022) argue that human neuroscience has two realistic paths to reliable brain-behavior correlations. One is to maximize the sample size, and the other is to maximize the effect size through tightly targeted, within-subject designs[74]. In line with the first path, large multi-site work by Marek and colleagues (2022) has shown that correlations between whole-brain MRI measures and complex traits are much smaller than many earlier single-site papers suggested. Additionally, replicable effects generally only emerge when sample sizes reach into the thousands. The study demonstrates that sample sizes in the tens or low hundreds tend to produce unstable effect sizes and poor replication[75]. From this perspective, having roughly 100 participants per cohort, the current study is well-powered by traditional imaging standards, but clearly underpowered for BWAS-scale effects. Under that assumption, developed CPM models trained on 100 individuals were unlikely to uncover a generalizable rsFC-behavior mapping, and the failure to predict SRP-SF total or factor scores above a mean baseline is explanatory. The second path outlined by Gratton et. al. is to maximize effect sizes through focused, high-precision investigations. Although the present study already targets a relatively specific phenotype in a community sample, the null findings suggest that rsFC-psycopathy associations are heterogeneous and not captured by a uniform connectivity pattern. This possibility motivated the use of IS-RSA, which is designed to target the brain-behavior relationships at the level of subject-to-subject similarity.

Finally, there are methodological reasons why CPM, as implemented in this study, may struggle to capture the brain-behavior relationships. CPM assumes a linear, additive mapping from edge weights to trait scores: each selected connection contributes independently and with a constant slope[26]. If the actual mapping is non-linear, depends on interactions between edges, or differs across groups, a linear model may not well approximate such relationships. The null CPM findings observed in this thesis underline the importance of future work investigating complementary non-linear approaches for prediction.

### 5.3 Post-hoc analyses

The null results obtained during prediction motivated us to explore the heterogeneity of the Japanese sample by using inter-subject representational similarity analysis. The nearest-neighbor model assumes that subjects close in the psychopathy continuum have similar functional connectivity patterns at particular regions. In a heterogeneous construct like psychopathy, where people with the same total score can have very different profiles (e.g., low on affective and high on interpersonal facets, and vice versa), this is a strong assumption. This study empirically shows that simply being close in total score does not imply closeness in the connectivity space.

Across the three IS-RSA variants, which each operationalized behavioral similarity in a different way, the clearest pattern emerged from the AnnaK model. This approach tests whether individuals with higher psychopathy scores show a more similar connectivity pattern to one another than would be expected by chance. In these post-hoc analyses, network enrichment suggested an overrepresentation of default mode network edges, meaning that individuals at the higher end of psychopathy continuum share a slightly more similar DMN-centered organization compared to lower- or mid-scoring individuals. This finding broadly aligns with a recent resting-state study that combines person-specific network modelling with dimensional psychopathy measures. Dotterer et al. (2020) emphasize that the neural networks underlying psychopathic traits in young men are highly individualized overall, yet individuals with higher affective psychopathy features show increased density of connections between DMN and central executive networks[76].

A complementary perspective comes from the item-wise IS-RSA model, in which behavioral similarity was computed from the whole pattern of item responses rather than a single total score. Two individuals are considered similar if they endorse a similar configuration of responses across all SRP-SF questions, even if their overall scores differ. Interestingly, the model produced weak effects in default mode, salience, and control networks, and the resulting map showed the clearest correspondence with the “psychopathy network”(Dugré and De Brito, 2025[19]). The fact that item-wise IS-RSA yielded these results, where the NN model did not, could suggest that profile-level information across the spectrum carries a meaningful neural signal that is “washed out” when psychopathy is reduced to a single score. This fits with the broader argument that heterogeneity in psychopathy matters. Latzman et al.’s commentary on Poeppl et al.[77], for instance, explicitly warns that interpersonal-affective and impulsive-antisocial components show contrasting, sometimes opposing associations with behavioral and brain-based outcomes, and that global scores can obscure these effects[78].

### 5.4 Importance of considering culture

The replicability of null findings across culturally distinct samples highlights another key limitation of neuroimaging research. Most of what is currently known about the neural correlates of psychopathology comes from WEIRD samples[79]. At the same time, the cultural neuroscience literature shows that culture can reliably modulate neural responses in social and affective tasks[80–82]. Although a small number of neuroimaging studies have now examined psychopathic traits in non-Western samples[30, 83–86], there is, to our knowledge, no systematic cross-cultural neuroimaging literature on psychopathy. As a result, there is no strong empirical basis to assume that psychopathy-brain mappings are identical across cultures. In this context, the present study contributes a modest first step by including two community samples from different cultural backgrounds (Japan and Germany). Both samples are drawn from relatively high-functioning, high-income populations, but they differ in cultural norms and social context. These converging null results suggest that the main obstacle to predictive modelling might not be primarily cultural. Instead, the challenges identified earlier - very small effect sizes, heterogeneity of psychopathy as a phenotype, and the limitations of linear models - appear to apply broadly in similar ways to both Japanese and German adults. Taken together, these points highlight that, given our current state of knowledge, the primary challenge is to identify reliable neural correlates of psychopathic traits on a continuum before we can address finer-grained questions about cultural modulation.

### 5.5 Limitations

Several limitations of the present work should be acknowledged when interpreting the findings. First, although the sample sizes in both cohorts are typical for fMRI studies, large-scale analyses of brain-behavior relationships indicate that typical correlations between whole-brain functional connectivity and complex traits often require thousands of participants for stable, reproducible effects[74, 75]. Second, the CPM framework[26] used in the study also imposed several constraints. Following the original approach, CPM was implemented as a linear, additive model that does not take into consideration that brain-psychopathy relationships could be non-linear or could rely on higher-order interactions. Additionally, although the IS-RSA maps were aligned with prior network-level findings, most effects did not survive the correction for multiple comparisons and should therefore be considered with caution. Third, both samples used included individuals similar in socio-economic context. Thus, the results might not extend to culturally and socio-economically diverse groups. The results are also not extendable to adolescents or clinical/forensic populations with more extreme psychopathy scores. Finally, researchers must remain critical regarding the phenotyping of psychopathic traits, since both cohorts relied on self-report instruments (SRP-SF in Japan, SD3 psychopathy subscale in Germany), even though the evidence suggests that self-report questionnaires are a valid and reliable method for measuring psychopathic traits.

### 5.6 Future directions

These limitations point towards several promising avenues for future work. First, there is a need to study larger, richer, and more diverse samples. Multi-site datasets with thousands of participants, harmonised psychopathy measures, and coverage spanning community, clinical, and forensic samples would allow the field to move towards replicable and robust predictive models. The ENIGMA Antisocial Behavior Working Group is one prominent initiative pursuing exactly this kind of large-scale, harmonized approach (ENIGMA-ASB; Thompson et al., 2020[87]). Second, considering the uncovered heterogeneity of psychopathy, future research would also benefit from individualized analyses. Specifically, working in the context of precision psychiatry, person-specific brain mapping could reveal functional estimates that are much more reliable and reveal robust features of network organization that are not visible in group-level data[88, 89]. Third, relatedly, functional connectome fingerprinting studies demonstrate that individuals can be reliably identified from their connectivity patterns alone[90], underscoring the strength of person-specific signatures in the functional connectome. Thus, incorporating within-individual analyses into studies of psychopathic traits may help to reveal meaningful individual differences in functional connectivity and to pinpoint neural correlates of psychopathy on an individual level.

Taken together, the results of this thesis suggest that resting-state functional connectivity does not currently provide a robust, generalizable predictive marker of psychopathic traits in community adults, at least in the Japanese and German samples studied here. Nonetheless, across CPM, IS-RSA and network-enrichment analyses, a picture emerged in which default mode, salience and frontoparietal networks appeared to carry the possible signal related to psychopathy, somewhat in line with previous literature. Methodologically, by combining connectome-based predictive modelling and inter-subject representational similarity approaches, and by reporting both null and exploratory findings, this thesis illustrates the limitations of resting-state functional connectivity as a predictive marker in psychopathy research. It also highlights the need to account for individual heterogeneity across the continuum and to move towards more individual-centered approaches as the next necessary step.

## 6 Conclusion

The present study was preregistered and conducted in line with current open-science recommendations (link). The thesis aimed to test whether whole-brain resting-state functional connectivity can account for individual differences in psychopathic traits in a community and whether such relationships generalize across two culturally distinct samples. Across both Japanese and German cohorts, resting-state functional connectivity did not yield a robust, generalizable predictive marker for psychopathy total, factor, and facets scores. At the same time, inter-subject similarity analyses suggested that connectivity profiles - in particular within default mode, salience, and frontoparietal control networks - are highly heterogeneous among individuals at the lower end of the psychopathy scale, which likely limits the ability of simple linear models to capture robust brain-behavior relationships. However, the results also showed that in community adults, the associations are null and not easily converted into accurate individual-level predictions. Finally, the thesis highlights both the value and the limitations of the utilized approaches. Predictive modelling can still uncover meaningful patterns in resting-state connectivity in relation to psychopathic traits, but the present results underscore the need for larger and more diverse samples, possibly more fine-grained psychopathy phenotyping, and more individual-centered frameworks. Together, this work clarifies what resting-state connectivity can realistically offer for predictive modelling and points towards more precision-oriented directions for future research.

## Data Availability

All data produced in the present study are available upon reasonable request to the authors.

## Code Availability

Baseline Connectome Predictive Modelling was conducted using scripts from Medical Machine Learning Lab - University of Mü nster: https://github.com/wwu-mmll/confound_corrected_cpm. Weighted and NBS CPM models were implemented based on the original CPM code from the same repository. Inter-subject representational similarity analyses were performed based on the IS-RSA Tutorial by Emily Finn and Luke Chang (https://naturalistic-data.org/content/Intersubject_RSA.html). Customized code for all analyses is available at request.

## Notes

### Competing Interest Statement

The authors have declared no competing interest.

### Funding Statement

This study did not receive any funding.

### Author Declarations

Ethics Committee of Kyoto University gave ethical approval for this work. Ethics Committee at the medical faculty of the University of Leipzig gave ethical approval for this work.

### Summary of Updates

This version of the manuscript has been revised to update attributions in the Introduction, specifically references to primary sources on two-factor and four-facet models of psychopathy. No changes to analyses or results.

